# Machine learning for improved dengue diagnosis, Puerto Rico

**DOI:** 10.1101/2024.11.13.24317272

**Authors:** Zachary J. Madewell, Dania M. Rodriguez, Maile B. Thayer, Vanessa Rivera-Amill, Jomil Torres Aponte, Melissa Marzan-Rodriguez, Gabriela Paz-Bailey, Laura E. Adams, Joshua M. Wong

## Abstract

**Background:** Diagnosing dengue accurately, especially in resource-limited settings, remains challenging due to overlapping symptoms with other febrile illnesses and limitations of current diagnostic methods. This study aimed to develop machine learning (ML) models that leverage readily available clinical data to improve diagnostic accuracy for dengue, potentially offering a more accessible and rapid diagnostic tool for healthcare providers.

**Methods:** We used data from the Sentinel Enhanced Dengue Surveillance System (SEDSS) in Puerto Rico (May 2012—June 2024). SEDSS primarily targets acute febrile illness but also includes cases with other symptoms during outbreaks (e.g., Zika and COVID-19). ML models (logistic regression, random forest, support vector machine, artificial neural network, adaptive boosting, light gradient boosting machine [LightGBM], and extreme gradient boosting [XGBoost]) were evaluated across different feature sets, including demographic, clinical, laboratory, and epidemiological variables. Model performance was assessed using the area under the receiver operating characteristic curve (AUC), where higher AUC values indicate better performance in distinguishing dengue cases from non-dengue cases.

**Results:** Among 49,679 patients in SEDSS, 1,640 laboratory-confirmed dengue cases were identified.□The□XGBoost and LightGBM models achieved the highest diagnostic accuracy, with AUCs exceeding 90%, particularly with comprehensive feature sets. Incorporating predictors such as monthly dengue incidence, leukopenia, thrombocytopenia, rash, age, and absence of nasal discharge significantly enhanced model sensitivity and specificity for diagnosing dengue. Adding more relevant clinical and epidemiological features consistently improved the models’ ability to correctly identify dengue cases.

**Conclusions:** ML models, especially XGBoost and LightGBM, show promise for improving diagnostic accuracy for dengue using widely accessible clinical data, even in resource-limited settings. Future research should focus on developing user-friendly tools, such as mobile apps, web-based platforms, or clinical decision systems integrated into electronic health records, to implement these models in clinical practice and exploring their application for predicting dengue.

**Author summary:** Dengue is a tropical disease caused by the dengue virus, which is transmitted by mosquitoes. It affects millions of people worldwide every year, leading to severe illness and even death in some cases. Accurate and timely diagnosis of dengue is crucial for proper treatment and controlling the spread of the virus. Traditionally, diagnosing dengue relies on symptoms and laboratory tests, which can sometimes be non-specific and not immediately available in distinguishing dengue from other similar illnesses. In our study, we explored the use of machine learning, a type of artificial intelligence, to improve dengue diagnosis using patient information from Puerto Rico. Our models, which use information like age, symptoms, and specific blood cell counts, can accurately predict whether someone has dengue. We found that some simple information, like whether a patient has a rash or low blood cell counts, can be very helpful in making a diagnosis. While more complex models performed slightly better, simpler models can also be effective, especially in places with limited resources. Our study shows that using computer models can improve dengue diagnosis and help healthcare providers make better decisions for their patients.

## Introduction

Dengue, a mosquito-borne viral infection transmitted by *Aedes* mosquitoes, is a significant global public health threat. Endemic in over 100 countries and affecting 2.5 billion people at risk, the virus poses a substantial burden on healthcare systems worldwide [1–3]. Dengue is caused by four distinct dengue viruses, also known as serotypes (DENV-1, -2, -3, and -4) that can lead to a spectrum of clinical presentations. These range from asymptomatic infections to debilitating illness, and in severe cases, potentially life-threatening complications like dengue hemorrhagic fever and dengue shock syndrome [4–6]. Globally, dengue is estimated to cause 390 million infections [7] and 40,500 deaths annually [8].

The United States unincorporated territory of Puerto Rico has a long history with dengue, with the first reported outbreak in 1899 [9]. Puerto Rico accounts for over 95% of all locally acquired dengue cases reported within the United States [10, 11]. Transmission follows a seasonal pattern, with large outbreaks typically occurring every three to five years [12]. From 2010–2020, dengue was associated with nearly 30,000 confirmed and probable cases including 584 severe cases, 10,000 hospitalizations, and 68 deaths island-wide [11]. A large epidemic in Puerto Rico occurred in 2012–2013, dominated by DENV-1, although sporadic cases of DENV-4 also occurred. During 2016–2023, cases remained relatively low compared to historical levels. However, in early 2024, a concerning surge in cases prompted a public health emergency declaration by Puerto Rico’s Department of Health. The outbreak has placed increased pressure on the healthcare system, which was already strained by the ongoing syndemic of dengue, influenza, and COVID-19 [13]. Particularly, high bed occupancy observed in June 2024 due to COVID-19 has underscored the urgent need for improved diagnostic methods and effective public health interventions to manage and control the spread of dengue and other co-circulating pathogens.

Diagnosing dengue accurately can be difficult, particularly in the early stages of the illness, due to overlapping symptoms with other febrile illnesses and the need for repeated evaluations to distinguish dengue from other conditions as the illness progresses. Current diagnostic methods have their limitations as well. Laboratory tests for dengue, such as RT-PCR or serologic assays, require specialized equipment and expertise, are often unavailable in resource-limited settings, and can take several days to return results, making them less suitable for guiding immediate clinical management decisions. These tests also have limited windows of detection; for example, PCR is most sensitive in the first week of illness, while serologic tests are more useful later, potentially missing cases depending on when patients present for care. Rapid tests, while faster, have limitations in sensitivity, can be costly, and have not been authorized for use in the United States. As a result, many dengue cases are not confirmed in time to guide clinical decisions, which can impact patient care, public health efforts, and accurate reporting of disease burden. For example, during outbreaks, many cases are clinically diagnosed based on symptoms and epidemiological context, as laboratory resources are often prioritized for severe cases or sentinel surveillance rather than confirming every suspected case.

This project proposes a novel approach to address these challenges by developing machine learning (ML) models for diagnosing dengue fever. These models leverage readily available data from the Sentinel Enhanced Dengue Surveillance System (SEDSS) in Puerto Rico, including demographics, symptoms, laboratory, and other factors. The models could be integrated into existing electronic health records (EHR) systems to assist clinicians in making timely diagnoses [14]. In clinical practice, the ML model could automatically analyze patient data upon entry, generating a risk score for dengue based on the combination of symptoms, patient history, and other available data [15]. When the model identifies a high probability of dengue, it would alert the clinician through the EHR system, prompting further investigation or specific diagnostic testing. This approach has been successfully used in other diseases, such as COVID-19 [16], infective endocarditis [17], and incident atrial fibrillation [18], where ML models are embedded into EHRs to provide early warnings for timely interventions, improving patient outcomes.

In resource-limited settings where EHR systems may not be universally available, these models could be adapted for use on portable devices like smartphones or tablets, enabling healthcare workers to input patient data and receive a risk score even when access to advanced diagnostic tools is limited. This flexibility ensures that the benefits of ML-driven diagnostic support can be extended beyond well-resourced environments.

By tapping into the growing adoption of EHRs worldwide, ML models can offer a solution that is not constrained by the physical limitations of laboratory resources. In many regions, even where basic laboratory facilities are lacking, EHR systems are being rapidly adopted, allowing healthcare providers to store and analyze patient data electronically. This technological advancement presents a unique opportunity for ML to enhance diagnostic accuracy and provide timely insights using data that is already being collected. To identify the optimal approach, we compare the performance of multiple ML algorithms and sampling techniques alongside traditional logistic regression analysis. By using ML, this project aims to improve diagnostic accuracy for dengue compared to traditional methods, provide a rapid and accessible tool for resource-limited settings, and potentially reduce misdiagnosis rates to improve patient outcomes. This project aligns with the growing body of research exploring the use of ML for infectious disease diagnosis, particularly in areas with limited diagnostic resources [19–22].

## Methods

### Study population

In this analysis, we used data from SEDSS, an ongoing facility-based study in Puerto Rico that tracks the frequency and causes of acute febrile illness [23, 24]. Since its inception in 2012, SEDSS has included five sites: 1) Saint Luke’s Episcopal Hospital (SLEH) in Ponce, a tertiary acute care facility (2012–present), 2) SLEH-Guayama, a secondary acute care hospital (2013–2015), 3) Hospital de La Universidad de Puerto Rico in Carolina, another secondary acute care teaching hospital (2013–2015), 4) Centro de Emergencia y Medicina Integrada (CEMI), an outpatient acute care clinic in Ponce (2016– present), and 5) Auxilio Mutuo Hospital, a tertiary care facility in the San Juan Metro Area (2018– present). The data used for this analysis was downloaded from SEDSS on July 25, 2024.

### Study enrollment and data collection

Participants in SEDSS are enrolled through convenience sampling. Potential participants are identified by triage nurses as any patient with an acute febrile illness (AFI) defined by the presence of fever (≥38.0°C for temperatures measured orally, ≥37.5°C for temperatures measured rectally, and ≥38.5°C for temperatures measured axillary) at the time of triage or chief complaint of having a fever within the past 7 days. During the Zika epidemic in Puerto Rico (June 2016–June 2018), patients were eligible if they presented with either rash and conjunctivitis, rash and arthralgia, or fever [25]. Starting in April 2020, patients with cough or dyspnea within the last 14 days (with or without fever) were also eligible to better capture respiratory viruses [13]. No age groups were excluded, although infants were only eligible for enrollment if they presented to the hospital after their initial discharge after birth. After meeting the inclusion criteria and being informed about the study, participants provided written informed consent. In cases where patients were incapacitated at the time of triage due to acute illness, consent was sought after their stabilization.

Data collection in SEDSS involves patient interviews and medical record reviews at both enrollment and convalescence (∼7–14 days later). The case investigation form (CIF) gathers demographics, comorbidities, and clinical features. The convalescent sample processing form (CSPF) echoes CIF data, adding the second specimen collection date and AFI severity indicators (hospitalizations, clinic visits). Initially paper-based (CIF/CSPF), data collection transitioned to electronic format in 2020 using REDCap on Android tablets [26, 27].

### Sample collection and laboratory procedures

Blood, nasopharyngeal (NP), and oropharyngeal (OP) specimens were collected at enrollment from eligible participants. Additional blood samples (serum and whole blood) were also collected during the convalescent phase. Participation required providing at least one sample (blood or OP/NP swab). All patients had molecular testing for dengue virus for specimens collected within 7 days of symptom onset. Serologic testing for anti-DENV antibodies was performed using Immunoglobulin M (IgM) antibody capture enzyme-linked immunosorbent assay (ELISA) for specimens collected >3 days after symptom onset (6). Dengue cases were defined as a positive result for DENV by RT-PCR or DENV IgM.

### Variable selection

Our variable selection process was designed to identify features at the initial clinical presentation that could assist in accurately diagnosing laboratory-confirmed dengue cases. We began by considering 54 variables based on physicians’ medical knowledge and clinical experience. These variables included demographic characteristics, recent travel history, warning signs, other clinical signs, and laboratory findings. Additionally, dengue monthly incidence in Puerto Rico during 2012 to 2024 was obtained from the Puerto Rico Passive Arboviral Disease Surveillance System (PADSS). This data informed prior knowledge that could influence a physician’s dengue diagnosis, consistent with previous research [21].

To refine feature selection, we used logistic regression to calculate crude unadjusted odds ratios for each of the 54 variables. The use of unadjusted odds ratios allowed us to quickly assess the strength of association between each variable and dengue diagnosis without the complexity of adjusting for confounders at this stage. This step helped to screen variables and group them into feature sets based on their association with dengue, as indicated by the magnitude of their odds ratios. The rationale for this approach is that, in clinical practice, certain variables with strong associations (e.g., high or low odds ratios) may be more immediately relevant for consideration in a diagnostic model. This is particularly useful in scenarios where we want to simplify the feature selection process while still capturing the most impactful predictors. By using different odds ratio cutoffs (e.g., >6 or <0.17, >3 or <0.33, >2 or <0.50), we created four groups of variables with varying levels of association strength. Additionally, we included a group of variables that were statistically significant at p<0.05, to ensure that we did not overlook potentially important features with more moderate associations.

The decision to use these crude odds ratios to define feature sets was driven by the need for a straightforward and clinically intuitive method to narrow down the initial list of 54 variables before applying more complex ML models. Although ML algorithms can automatically select and weight features during model training, this initial step allowed us to focus on variables that were already known to be clinically relevant or strongly associated with dengue, simplifying the interpretation of the models for clinicians. The final feature sets were then evaluated using various ML models to determine their predictive performance. The rationale for evaluating different sized feature sets was to understand how model performance might change as we included more or fewer variables. This approach helped us balance the trade-off between model complexity and predictive accuracy, particularly when considering the potential application of these models in different clinical settings.

Although previous dengue infection is a known risk factor for developing severe disease and can influence clinical presentation, it was excluded from the primary analysis due to limitations in its applicability to routine clinical practice. Immunoglobulin G (IgG) testing is not universally available, and interpreting positive IgG results can be challenging for clinicians, potentially leading to misdiagnosis. However, to assess the potential impact of this variable on model performance, we conducted an additional analysis incorporating a history of prior dengue infection (confirmed by IgG) as an additional feature in the highest performing model within each feature set. To further explore the performance of the models in resource-constrained settings, where complete blood counts (CBCs) might not be readily available, we conducted an additional subanalysis excluding leukopenia and thrombocytopenia from the highest performing models within each feature set.

### Sampling

To address class imbalance in our dataset, we used downsampling to ensure an equal representation of dengue-positive and dengue-negative cases [28]. Class imbalance, where dengue-positive cases are significantly outnumbered by dengue-negative cases, can bias ML models towards the majority class, reducing their ability to accurately predict minority class outcomes. Downsampling mitigates this issue by randomly reducing the number of majority class instances to match the number of minority class instances, thus balancing the dataset and improving the model’s ability to accurately classify dengue-positive cases. This process was performed using the downSample function from the caret package in R [29], setting the ratio of positive to negative cases to 1:1. The datasets were then randomly partitioned into training and testing sets using a 70:30 ratio. The training sets were used to train the ML models, whereas the testing set was reserved for model evaluation.

### Machine learning models

We used an initial logistic regression model as a baseline to explore the relationship between potential predictors and dengue diagnosis outcomes. This model was trained using the same groups of variables based on different odds ratio cutoffs as mentioned earlier: variables with odds ratios >6, >3, >2, and any variables significant at p<0.05. Stepwise selection was used to iteratively add or remove variables to identify the optimal model with the lowest Akaike Information Criterion (AIC). This approach balances model complexity and goodness-of-fit by selecting variables that contribute significantly to the model. The final logistic regression model, derived from stepwise selection, was evaluated on both the training and testing sets.

Six ML methods were used to predict dengue infections and assess feature importance. These algorithms were selected to represent a breadth of ML predictive models and reflect those commonly used as diagnostic tools for various diseases. The algorithms used include Random Forest (RF), Support Vector Machine (SVM), Artificial Neural Network (ANN), Adaptive Boosting (AdaBoost), Light Gradient Boosting Machine (LightGBM), and eXtreme Gradient Boosting (XGBoost). RF is an ensemble learning method that constructs multiple decision trees during training and outputs the mode of their predictions for classification or the mean prediction for regression [30]. SVM is a supervised learning algorithm that finds the optimal hyperplane to separate data points of different classes in the feature space [31]. ANN is a computational model inspired by the way biological neural networks work, consisting of interconnected nodes (neurons) that process input data to learn and make predictions [32]. AdaBoosting is an ensemble technique that combines the predictions of several weak classifiers to create a strong classifier by focusing more on the instances that previous classifiers misclassified [33]. LightGBM is a fast, distributed, high-performance gradient boosting framework that uses tree-based learning algorithms for efficient and accurate predictions [34]. XGBoost is an optimized gradient boosting framework that uses decision trees and a range of enhancements for improved performance and speed in predictive modeling [34]. In this study, we specifically used the “gbtree” booster, which implements decision tree-based gradient boosting for learning the model.

For parameter optimization, we used a grid search to evaluate different combinations of hyperparameter values, and the area under the receiver operating characteristic curve (AUC-ROC) was used as the optimization metric. We used 5-fold cross-validation during model training to ensure robustness and reduce overfitting. Specific details of the grid search strategy and parameters included in each model are provided in the Table S2. For gradient boosting models, the training process used 100 boosting rounds with early stopping if the evaluation metric did not improve for 10 rounds. The R packages randomForest [35], e1071 [36], nnet [37], ada [38], lightgbm [39], and xgboost [40], were used for implementing RF, SVM, ANN, AdaBoost, LightGBM, and XGBoost models, respectively.

### Performance evaluation

Model performance was evaluated on both the training and testing sets using AUC-ROC as the primary performance metric. AUC-ROC is an aggregate measure of performance across all possible classification thresholds, providing a comprehensive assessment of the model’s ability to distinguish between classes. We used the DeLong method to calculate the confidence intervals for the AUC-ROC to ensure accurate estimation of the model’s performance [41]. Additionally, confusion matrices were generated to assess the model’s classification performance in terms of sensitivity, specificity, and accuracy. The optimal threshold for classification was determined using Youden’s index from the ROC curve [42]. Cohen’s kappa was also calculated to measure the agreement between predicted and observed classifications, accounting for chance agreement.

Feature importance was assessed for each ML model. By quantifying each feature’s contribution to the model’s accuracy, we can identify the most influential features to better understand and improv predictions for each method. For RF, AdaBoost, LightGBM, and XGBoost, we used the varImp, varplot, lgb.importance, and xgb.importance functions from the randomForest [35], ada [38], lightgbm [39], and xgboost [40] packages. For SVM, we determined feature importance by calculating the absolute value of the coefficients from the support vectors. For ANN, we used permutation importance [43], which works by calculating the original AUC-ROC as a baseline, then permuting each feature to break its relationship with the target variable and recalculating the AUC-ROC. The importance of a feature is determined by the decrease in AUC-ROC after permutation; a significant drop indicates high importance. Predicted probabilities were uniformly binned, with mean predicted probabilities and observed positive case proportions calculated for each bin. All analyses were done using R version 4.4.0 [44].

### Post-hoc subanalysis: Evaluating AUC with sequential feature addition

After identifying feature sets based on crude odds ratios in the primary analysis, we performed a post-hoc subanalysis to evaluate model performance by sequentially adding the most important features from the highest-performing ML model. This approach simulates a refined diagnostic process where key features are known in advance. By recalculating AUC with each feature added, we assessed the incremental benefit of each and identified the minimum set of variables for optimal performance. This analysis also tested the robustness of our primary feature selection by comparing it to ML-derived feature importance.

### Ethics statement

The Institutional Review Boards at the Centers for Disease Control and Prevention (CDC), Auxilio Mutuo, and Ponce Medical School Foundation approved the SEDSS study protocols 6214, and 120308-VR/2311173707, respectively. Written consent to participate was obtained from all adult participants and emancipated minors. For minors aged 14 to 20 years, written consent was obtained, and for those aged 7 to 13 years, parental written consent and participant assent were obtained.

## Results

### Participant Characteristics

From May 2012 to June 2024, 51,219 unique AFI visits were recorded from 41,180 participants enrolled in SEDSS, including 8,035 hospitalizations or transfers and 73 deaths. Of these visits, there were 49,679 AFI visits from 40,124 participants tested for DENV. The median age among was 15 years (interquartile range (IQR): 4, 38) and 52.8% were female (Table 1). From these, 1,640 (3.3%) had dengue (1,167 positive by RT-PCR, 473 positive by DENV IgM), 1,026 (62.5%) of which occurred during the 2012–2013 epidemic. SEDSS participants tested positive for other arboviruses including chikungunya (n=2,291) and Zika (n=1,899), and respiratory viruses including influenza A (n=4,296), influenza B (n=1,726), human adenovirus (n=1,835), respiratory syncytial virus (n=1,576), and SARS-CoV-2 (n=2,322), among others (S3 Table). The majority of the 1,155 serotyped dengue cases were DENV-1 (n=903, 78.2%), followed by DENV-3 (n=111, 9.6%), DENV-2 (n=91, 7.9%), and DENV-4 (n=50, 4.3%). Of 1,640 dengue cases, 737 (44.9%) were hospitalized or transferred, and two (0.1%) died. Median duration from symptom onset to presentation at the emergency room was 3 days [IQR: 1, 4] for dengue cases. A previous dengue infection, as indicated by a positive IgG test on or before the fifth day of illness, was present in 79.1% (n=564/713) of dengue cases tested. Additionally, 38.5% (n=632) had at least one comorbidity, including obesity (28.4%), chronic pulmonary disease (20.2%), and hypertension (9.0%).

**Table 1.** Demographic and clinical characteristics of participants (dengue and non-dengue cases), SEDSS, May 2012–June 2024. Dengue cases had a positive result for dengue by RT-PCR or IgM.

|  | Total<br>N = 49679<br>n (%) | Laboratory-confirmed<br>dengue cases<br>N = 1640<br>n (%) | Non-dengue<br>cases<br>N = 48039<br>n (%) | p-value |
| --- | --- | --- | --- | --- |
| **Month** |  |  |  | <0.001 |
| January | 3706 (7.5) | 133 (8.1) | 3573 (7.4) |  |
| February | 3792 (7.6) | 102 (6.2) | 3690 (7.7) |  |
| March | 3905 (7.9) | 71 (4.3) | 3834 (8.0) |  |
| April | 3939 (7.9) | 78 (4.8) | 3861 (8.0) |  |
| May | 4291 (8.6) | 107 (6.5) | 4184 (8.7) |  |
| June | 4377 (8.8) | 157 (9.6) | 4220 (8.8) |  |
| July | 4317 (8.7) | 144 (8.8) | 4173 (8.7) |  |
| August | 4489 (9.0) | 171 (10.4) | 4318 (9.0) |  |
| September | 4366 (8.8) | 163 (9.9) | 4203 (8.7) |  |
| October | 4423 (8.9) | 162 (9.9) | 4261 (8.9) |  |
| November | 4184 (8.4) | 199 (12.1) | 3985 (8.3) |  |
| December | 3890 (7.8) | 153 (9.3) | 3737 (7.8) |  |
| **Days post onset** |  |  |  | <0.001 |
| 0 | 8363 (17.8) | 119 (7.9) | 8244 (18.2) |  |
| 1-3 | 30321 (64.6) | 699 (46.5) | 29622 (65.2) |  |
| 4-6 | 6587 (14.0) | 621 (41.3) | 5966 (13.1) |  |
| 7+ | 1633 (3.5) | 65 (4.3) | 1568 (3.5) |  |
| **Age group** |  |  |  | <0.001 |
| <1 | 3449 (6.9) | 31 (1.9) | 3418 (7.1) |  |
| 1-4 | 10539 (21.2) | 107 (6.5) | 10432 (21.7) |  |
| 5-9 | 6640 (13.4) | 234 (14.3) | 6406 (13.3) |  |
| 10-19 | 7394 (14.9) | 665 (40.5) | 6729 (14.0) |  |
| 20-29 | 5885 (11.8) | 211 (12.9) | 5674 (11.8) |  |
| 30-39 | 4046 (8.1) | 109 (6.6) | 3937 (8.2) |  |
| 40-49 | 3294 (6.6) | 88 (5.4) | 3206 (6.7) |  |
| 50+ | 8432 (17.0) | 195 (11.9) | 8237 (17.1) |  |
| **Female sex** | 26238 (52.8) | 761 (46.4) | 25477 (53.0) | <0.001 |
| **Health region** |  |  |  | <0.001 |
| Aguadilla | 44 (0.1) | 2 (0.1) | 42 (0.1) |  |
| Arecibo | 197 (0.4) | 4 (0.2) | 193 (0.4) |  |
| Bayamon | 1041 (2.1) | 26 (1.6) | 1015 (2.1) |  |
| Caguas | 588 (1.2) | 28 (1.7) | 560 (1.2) |  |
| Fajardo | 359 (0.7) | 16 (1.0) | 343 (0.7) |  |
| Mayaguez | 118 (0.2) | 6 (0.4) | 112 (0.2) |  |
| Metro | 6523 (13.1) | 352 (21.5) | 6171 (12.8) |  |
| Ponce | 40667 (81.9) | 1205 (73.5) | 39462 (82.1) |  |
| Unknown | 142 (0.3) | 1 (0.1) | 141 (0.3) |  |
| **Previous DENV infection<sup>a</sup> (N = 2062)** |  |  |  | 0.003 |
| Yes | 1703 (82.6) | 564 (79.1) | 1139 (84.4) |  |
| No | 359 (17.4) | 149 (20.9) | 210 (15.6) |  |
| **Traveled to another country in past two weeks** | 1501 (3.0) | 74 (4.5) | 1427 (3.0) | 0.001 |
| **Household member with dengue** | 619 (1.2) | 133 (8.1) | 486 (1.0) | <0.001 |
| **Reported mosquito bites in past month** | 8557 (17.2) | 624 (38.0) | 7933 (16.5) | <0.001 |
| **Comorbidities** |  |  |  |  |
| Chronic pulmonary disease or asthma | 9721 (19.6) | 331 (20.2) | 9390 (19.5) | 0.544 |
| Cancer | 907 (1.8) | 26 (1.6) | 881 (1.8) | 0.519 |
| Chronic kidney disease | 607 (1.2) | 12 (0.7) | 595 (1.2) | 0.085 |
| Congenital heart disease | 1822 (3.7) | 52 (3.2) | 1770 (3.7) | 0.307 |
| Diabetes | 4065 (8.2) | 105 (6.4) | 3960 (8.2) | 0.009 |
| High cholesterol | 3034 (6.1) | 80 (4.9) | 2954 (6.1) | 0.039 |
| Hypertension | 6664 (13.4) | 147 (9.0) | 6517 (13.6) | <0.001 |
| Arthritis | 1954 (3.9) | 22 (1.3) | 1932 (4.0) | <0.001 |
| Thyroid disease | 2980 (6.0) | 69 (4.2) | 2911 (6.1) | 0.002 |
| Obesity | 9221 (18.6) | 199 (12.1) | 9022 (18.8) | <0.001 |
| Gastritis | 531 (1.1) | 27 (1.6) | 504 (1.0) | 0.028 |
| **Warning signs<sup>b</sup>** |  |  |  |  |
| Persistent vomiting | 9401 (18.9) | 409 (24.9) | 8992 (18.7) | <0.001 |
| Abdominal pain | 17476 (35.2) | 972 (59.3) | 16504 (34.4) | <0.001 |
| Restlessness | 18500 (37.2) | 767 (46.8) | 17733 (36.9) | <0.001 |
| Clinical fluid accumulation | 2939 (5.9) | 87 (5.3) | 2852 (5.9) | 0.093 |
| Mucosal bleeding | 4276 (8.6) | 255 (15.5) | 4021 (8.4) | <0.001 |
| Hepatomegaly | 119 (0.2) | 44 (2.7) | 75 (0.2) | <0.001 |
| **Other clinical signs/symptoms** |  |  |  |  |
| Fever | 45090 (90.8) | 1631 (99.5) | 43459 (90.5) | <0.001 |
| Conjunctivitis | 1762 (3.5) | 133 (8.1) | 1629 (3.4) | <0.001 |
| Chills | 26690 (53.7) | 1309 (79.8) | 25381 (52.8) | <0.001 |
| Nausea | 23125 (46.5) | 1127 (68.7) | 21998 (45.8) | <0.001 |
| No appetite | 29972 (60.3) | 1282 (78.2) | 28690 (59.7) | <0.001 |
| Rash | 9268 (18.7) | 924 (56.3) | 8344 (17.4) | <0.001 |
| Yellow skin | 793 (1.6) | 68 (4.1) | 725 (1.5) | <0.001 |
| Itchy skin | 6574 (13.2) | 603 (36.8) | 5971 (12.4) | <0.001 |
| Headache | 29778 (59.9) | 1383 (84.3) | 28395 (59.1) | <0.001 |
| Eye pain | 14106 (28.4) | 985 (60.1) | 13121 (27.3) | <0.001 |
| Myalgia | 23983 (48.3) | 1221 (74.5) | 22762 (47.4) | <0.001 |
| Arthralgia | 19948 (40.2) | 1020 (62.2) | 18928 (39.4) | <0.001 |
| Tachypnea | 9965 (20.1) | 744 (45.4) | 9221 (19.2) | <0.001 |
| Back pain | 17666 (35.6) | 874 (53.3) | 16792 (35.0) | <0.001 |
| Calf pain | 12638 (25.4) | 605 (36.9) | 12033 (25.0) | <0.001 |
| Arthritis | 4155 (8.4) | 242 (14.8) | 3913 (8.1) | <0.001 |
| Nasal discharge | 30350 (61.1) | 531 (32.4) | 29819 (62.1) | <0.001 |
| Sore throat | 22632 (45.6) | 599 (36.5) | 22033 (45.9) | <0.001 |
| Cough | 33842 (68.1) | 702 (42.8) | 33140 (69.0) | <0.001 |
| Diarrhea | 13803 (27.8) | 709 (43.2) | 13094 (27.3) | <0.001 |
| Hypotension | 1327 (2.7) | 133 (8.1) | 1194 (2.5) | <0.001 |
| Narrow pulse pressure | 828 (1.7) | 39 (2.4) | 789 (1.6) | 0.284 |
| Seizure | 654 (1.3) | 28 (1.7) | 626 (1.3) | 0.156 |
| Capillary refill <3 | 2955 (5.9) | 99 (6.0) | 2856 (5.9) | 0.889 |
| Pale skin | 11276 (22.7) | 772 (47.1) | 10504 (21.9) | <0.001 |
| Blue lips | 734 (1.5) | 70 (4.3) | 664 (1.4) | <0.001 |
| **Laboratory** |  |  |  |  |
| Leukopenia | 3757 (7.6) | 968 (59.0) | 2789 (5.8) | <0.001 |
| Thrombocytopenia | 4811 (9.7) | 738 (45.0) | 4073 (8.5) | <0.001 |
| **Monthly dengue incidence per 1000 people** | 0.01 [0.00, 0.02] | 0.12 [0.04, 0.32] | 0.01 [0.00, 0.02] | <0.001 |

<sup>a</sup> Previous DENV infection was defined as a positive DENV IgG test result collected within 5 days of symptom onset.
<sup>b</sup> Hemoconcentration (an increase in hematocrit due to plasma loss) is a known warning sign for severe dengue but was excluded from this study, as it requires serial measurements that are challenging to obtain at initial patient contact. To maintain applicability for early, point-of-care diagnosis, we focused on predictors available at first presentation.

Among the 1,640 dengue cases, the most frequent symptoms were fever (99.5%, an inclusion criterion for SEDSS), headache (84.3%), and chills (79.8%) (Table 1). Common warning signs included abdominal pain (59.3%), restlessness (46.8%), and persistent vomiting (24.9%). Additionally, 59.0% of cases presented with leukopenia and 45.0% had thrombocytopenia.

### Variable Selection

We identified key demographic, clinical, and laboratory characteristics significantly associated with dengue infection (Table S1) and created feature sets with 8 (OR>6 or <0.17), 20 (OR>3 or <0.33), 32 (OR>2 or <0.50), and 48 (p<0.05) variables, allowing us to assess the impact of different feature combinations on model accuracy (Table S1). The 8-variable feature set included age group, days post onset, rash, fever, leukopenia, thrombocytopenia, hepatomegaly, and self-reported history of dengue in a household member from participant questionnaire. The 20-variable set was derived from the 8-variable set, adding reported recent mosquito bites, chills, itchy skin, headache, chronic arthritis, eye pain, myalgia, tachypnea, nasal discharge, hypotension, pale skin, and blue lips. The 32-variable set also included diagnosis month, abdominal pain, conjunctivitis, nausea, arthralgia, diarrhea, cough, yellow skin, no appetite, back pain, mucosal bleeding, and dengue monthly incidence in Puerto Rico. The 48-variable set also included sex, health region, recent travel, arthritis, sore throat, calf pain, gastritis, narrow pulse pressure, persistent vomiting, restlessness, hypertension, thyroid disease, chronic kidney disease, diabetes, high cholesterol, and obesity. Chronic pulmonary disease, cancer, congenital heart disease, clinical fluid accumulation, seizure, and capillary refill <3 seconds were not associated with dengue infection at p<0.05 and were not included in ML models.

### Performance evaluation

In addition to the multivariable LR, we used a diverse range of ML algorithms to predict the probability of dengue infection. These ML models offer the advantage of adapting to complex data patterns and potentially improving diagnostic accuracy.

The 8-variable feature set had the lowest estimated AUC values across all models, ranging from 85.2% to 87.1% on the test set (Fig 1, Fig 2, Table 2). As the feature set size increased, AUCs progressively improved. The 20-variable set achieved 89.0% ∼ 89.5% AUC, and the 32-variable set reached 91.3% ∼ 94.2% AUC. The 48-variable set showed minimal further improvement (91.3% ∼ 94.7%) compared to the 32-variable set.

**Fig 1.**
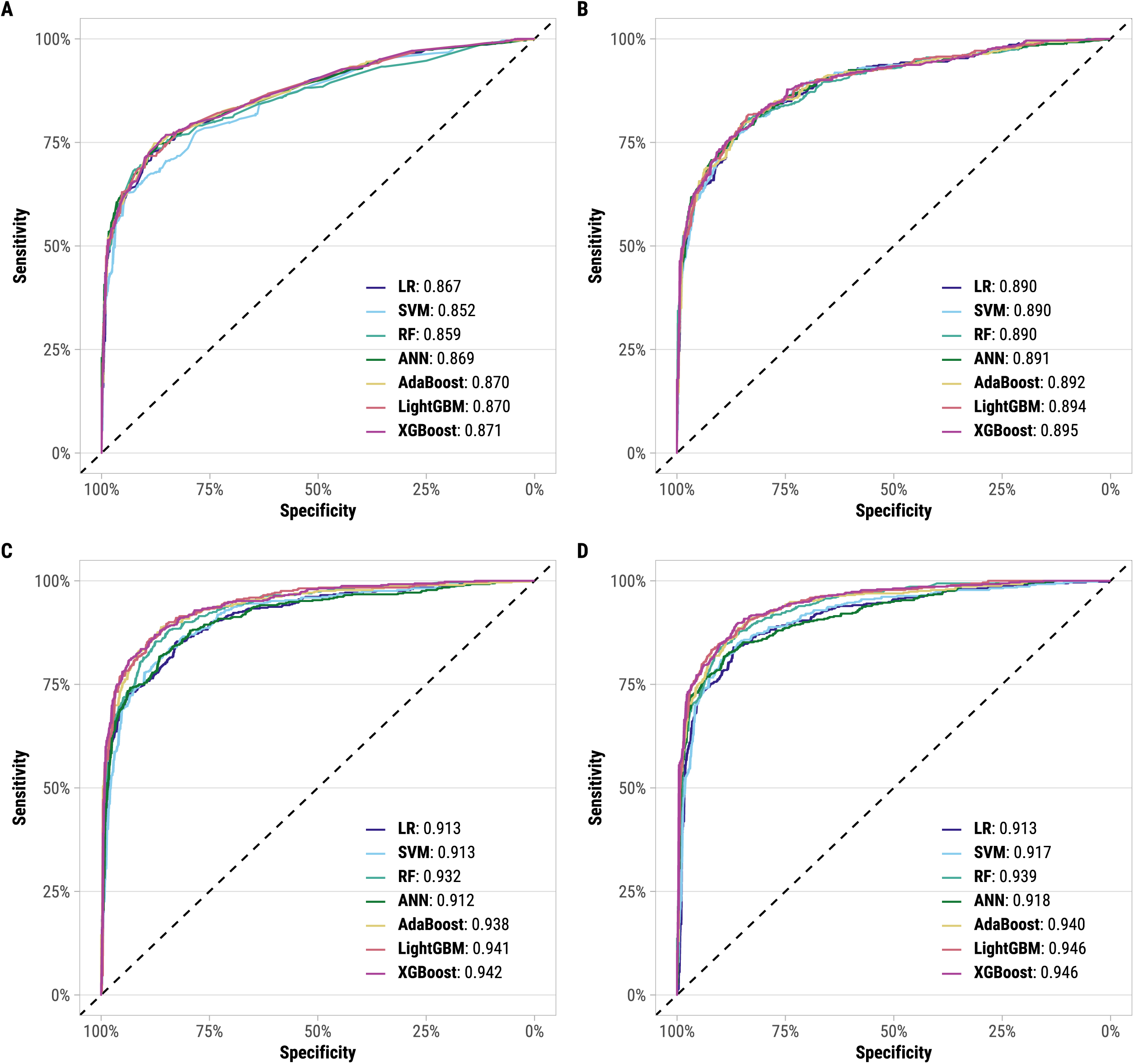
ROC curves of Logistic Regression, Support Vector Machine, Random Forest, Adaptive Boosting, Light Gradient Boosting, and Extreme Gradient Boosting models for (A) 8-, (B) 20-, (C) 32-, and (D) 48-variable feature sets, SEDSS, May 2012–June 2024. The area under the receiver operating characteristic curve is shown.

**Fig 2.**
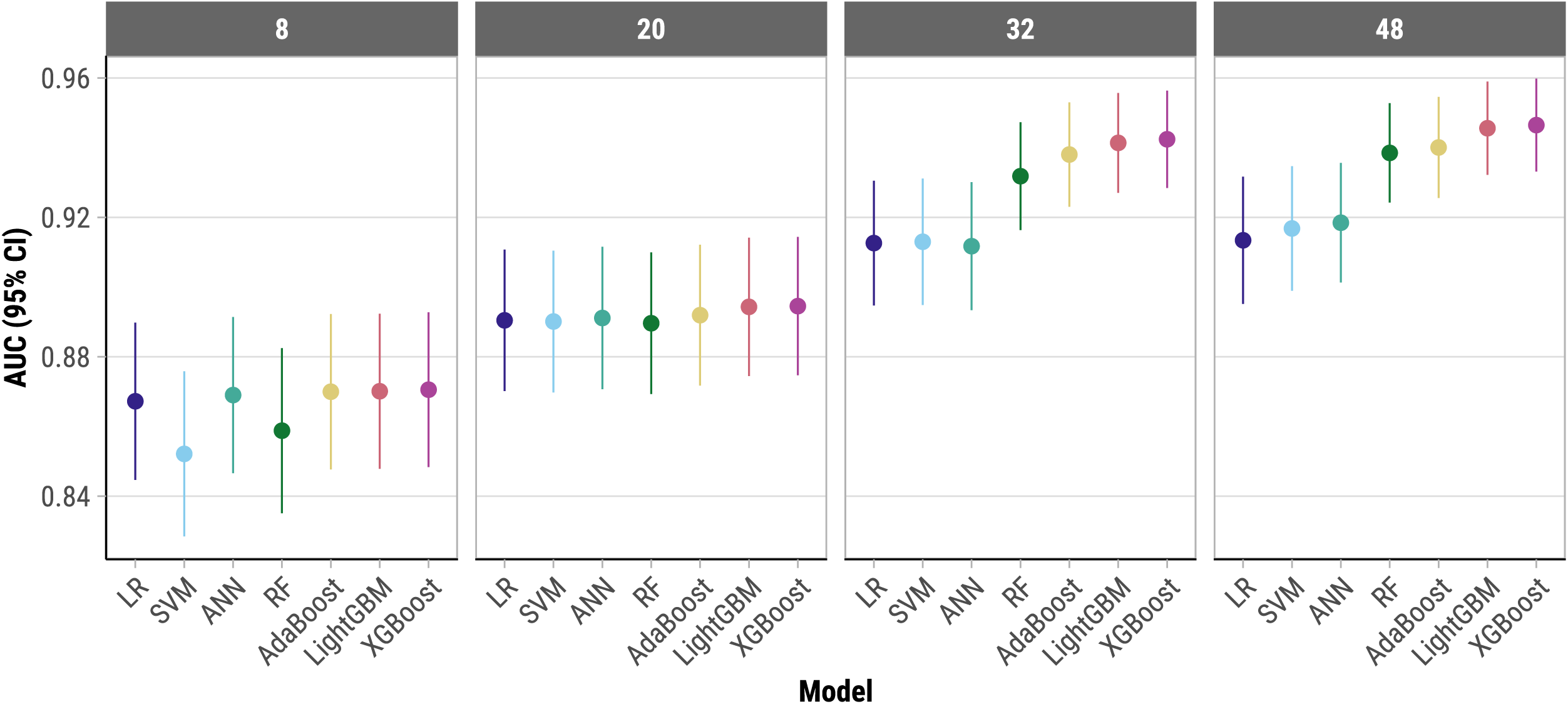
Forest plot of AUC values for Logistic Regression, Support Vector Machine, Random Forest, Adaptive Boosting, Light Gradient Boosting, and Extreme Gradient Boosting models for (A) 8-, (B) 20-, (C) 32-, and (D) 48-variable feature sets, SEDSS, May 2012–June 2024. DeLong method was used to obtain the 95% confidence intervals for the AUC-ROC.

**Table 2.** Performance of each algorithm for predicting dengue infection on the test set, SEDSS, May 2012–June 2024.

| Model | Accuracy | Sensitivity | Specificity | PPV | NPV | F1 Score | Kappa | AUC |
| --- | --- | --- | --- | --- | --- | --- | --- | --- |
| **8-variable feature set** |  |  |  |  |  |  |  |  |
| Logistic Regression | 0.8049 | 0.8821 | 0.7276 | 0.7641 | 0.8606 | 0.8189 | 0.6098 | 0.8672 |
| Support Vector Machine | 0.7734 | 0.8679 | 0.6789 | 0.7299 | 0.8371 | 0.7929 | 0.5467 | 0.8521 |
| Random Forest | 0.7998 | 0.9004 | 0.6992 | 0.7496 | 0.8753 | 0.8181 | 0.5996 | 0.8588 |
| Artificial Neural Network | 0.8089 | 0.8720 | 0.7459 | 0.7744 | 0.8535 | 0.8203 | 0.6179 | 0.8690 |
| Adaptive Boosting | 0.8089 | 0.8821 | 0.7358 | 0.7695 | 0.8619 | 0.8224 | 0.6179 | 0.8700 |
| Light Gradient Boosting | 0.8069 | 0.8963 | 0.7175 | 0.7603 | 0.8738 | 0.8228 | 0.6138 | 0.8701 |
| eXtreme Gradient Boosting | 0.8100 | 0.8516 | 0.7683 | 0.7861 | 0.8381 | 0.8176 | 0.6199 | 0.8706 |
| **20-variable feature set** |  |  |  |  |  |  |  |  |
| Logistic Regression | 0.8211 | 0.8476 | 0.7947 | 0.8050 | 0.8391 | 0.8257 | 0.6423 | 0.8905 |
| Support Vector Machine | 0.8079 | 0.8760 | 0.7398 | 0.7710 | 0.8565 | 0.8202 | 0.6159 | 0.8901 |
| Random Forest | 0.8120 | 0.8760 | 0.7480 | 0.7766 | 0.8578 | 0.8233 | 0.6240 | 0.8896 |
| Artificial Neural Network | 0.8191 | 0.8557 | 0.7825 | 0.7973 | 0.8443 | 0.8255 | 0.6382 | 0.8911 |
| Adaptive Boosting | 0.8211 | 0.8577 | 0.7846 | 0.7992 | 0.8465 | 0.8275 | 0.6423 | 0.8919 |
| Light Gradient Boosting | 0.8262 | 0.8374 | 0.8150 | 0.8191 | 0.8337 | 0.8281 | 0.6524 | 0.8943 |
| eXtreme Gradient Boosting | 0.8201 | 0.8780 | 0.7622 | 0.7869 | 0.8621 | 0.8300 | 0.6402 | 0.8945 |
| **32-variable feature set** |  |  |  |  |  |  |  |  |
| Logistic Regression | 0.8394 | 0.8272 | 0.8516 | 0.8479 | 0.8313 | 0.8374 | 0.6789 | 0.9126 |
| Support Vector Machine | 0.8343 | 0.8862 | 0.7825 | 0.8029 | 0.8730 | 0.8425 | 0.6687 | 0.9130 |
| Random Forest | 0.8628 | 0.8740 | 0.8516 | 0.8549 | 0.8711 | 0.8643 | 0.7256 | 0.9318 |
| Artificial Neural Network | 0.8415 | 0.8659 | 0.8171 | 0.8256 | 0.8590 | 0.8452 | 0.6829 | 0.9117 |
| Adaptive Boosting | 0.8648 | 0.8801 | 0.8496 | 0.8540 | 0.8763 | 0.8669 | 0.7297 | 0.9380 |
| Light Gradient Boosting | 0.8740 | 0.8882 | 0.8598 | 0.8636 | 0.8849 | 0.8758 | 0.7480 | 0.9414 |
| eXtreme Gradient Boosting | 0.8730 | 0.8760 | 0.8699 | 0.8707 | 0.8753 | 0.8734 | 0.7459 | 0.9424 |
| **48-variable feature set** |  |  |  |  |  |  |  |  |
| Logistic Regression | 0.8547 | 0.8699 | 0.8394 | 0.8442 | 0.8658 | 0.8569 | 0.7093 | 0.9134 |
| Support Vector Machine | 0.8547 | 0.8598 | 0.8496 | 0.8511 | 0.8583 | 0.8554 | 0.7093 | 0.9168 |
| Random Forest | 0.8659 | 0.8740 | 0.8577 | 0.8600 | 0.8719 | 0.8669 | 0.7317 | 0.9385 |
| Artificial Neural Network | 0.8547 | 0.8923 | 0.8171 | 0.8299 | 0.8835 | 0.8599 | 0.7093 | 0.9185 |
| Adaptive Boosting | 0.8730 | 0.8598 | 0.8862 | 0.8831 | 0.8634 | 0.8713 | 0.7459 | 0.9401 |
| Light Gradient Boosting | 0.8770 | 0.9207 | 0.8333 | 0.8467 | 0.9131 | 0.8822 | 0.7541 | 0.9456 |
| eXtreme Gradient Boosting | 0.8791 | 0.8618 | 0.8963 | 0.8926 | 0.8664 | 0.8769 | 0.7581 | 0.9465 |

PPV: positive predictive value; NPV: negative predictive value; AUC: area under receiver operating characteristic curve.

Among ML models, LightGBM and XGBoost consistently achieved the highest AUC across all feature sets, outperforming multivariable LR, SVM, and ANN by ∼3% for the 32- and 48-variable sets. This higher performance is likely due to their ability to efficiently handle large feature spaces, capture complex interactions, and prevent overfitting through advanced regularization techniques. Despite this, even simpler models like LR achieved strong performance with AUCs of 91% for the larger feature sets (32 and 48 variables), suggesting that core features captured a substantial amount of the predictive signal. AUC values exceeded 93% for all models in the 32- and 48-variable sets when evaluated on the training set (S4 Table).

All models had high sensitivity (>84%) across feature sets (Table 2). Specificity was lower for the 8-variable set (67.9 ∼ 76.8%) but improved to >85% for most models in the 32- and 48-variable sets. XGBoost with 48-variables achieved the highest overall AUC (94.7%), with corresponding sensitivity and specificity of 86.2% and 89.6%, respectively. Adding IgG testing improved the performance of XGBoost models across all feature sets, increasing AUC values by 2.6%, 2.0%, 0.5%, and 0.2% for the 8-, 20-, 32-, and 48-variable sets, respectively (S5 Table). Excluding leukopenia and thrombocytopenia from the XGBoost models resulted in a decrease in AUC values, ranging from 0.5% for the 48-variable set to 5.5% for the 8-variable set.

F1 scores consistently exceeded 82% across all models for the 20-, 32-, and 48-variable sets, indicating a good balance of precision (correct positive predictions out of all positive predictions) and recall (correct positive predictions out of all actual positives). Kappa values were highest (∼0.75) for LightGBM and XGBoost for the 32- and 48-variable feature sets, suggesting good agreement between model predictions and actual classifications, beyond what would be expected by chance.

### Feature importance

For the 8- and 20-variable sets, leukopenia was the top-performing feature for SVM, ANN, RF, LightGBM, XGBoost, and LR, followed by thrombocytopenia and rash (S1 Fig, S2 Fig, S6 Table). Age group, days post onset, eye pain, and absence of nasal discharge also emerged as important predictors for RF, LightGBM, and XGBoost. In contrast, AdaBoost identified blue lips, hepatomegaly, and hypotension as the highest scoring features. AdaBoost focuses on creating a series of weak learners (typically decision stumps) and combines them to form a strong classifier. It gives more weight to misclassified instances in subsequent iterations, which can result in a different feature importance profile as the model emphasizes different aspects of the data to improve accuracy. For the 32- and 48-variable sets, monthly dengue incidence showed the highest predictive power for SVM, RF, ANN, LightGBM, XGBoost, and LR (S3 Fig, S4 Fig).

In the subanalysis where the most important features identified by the highest performing ML model, XGBoost, were sequentially added to the feature set, we observed variable improvements in AUC values (S5 Fig). The initial addition of two variables increased the AUC by 2.8%, suggesting a substantial enhancement in model performance with the inclusion of high-priority predictors. As more variables were added, the gains in AUC became marginal, with incremental improvements ranging from 0% to 1.5%. By quantifying the gains in AUC through sequential addition of important features, this analysis highlights that a relatively small set of highly predictive features can achieve substantial diagnostic performance, reinforcing the utility of ML-based feature selection in optimizing clinical models.

## Discussion

The use of various ML algorithms, including RF, AdaBoost, LightGBM, and XGBoost, demonstrate significant improvements in diagnostic accuracy for dengue infection compared to traditional methods. Prior research has shown that the diagnostic accuracy of the 1997 and 2009 WHO clinical case definitions for dengue has high sensitivity (93%) but low specificity (29% and 31%, respectively), making it challenging to distinguish dengue from other febrile illnesses in clinical settings [45]. With AUC values exceeding 90% for larger feature sets, these models could potentially reduce misdiagnosis rates and improve patient outcomes, especially in settings where rapid tests are either not authorized or limited in availability. Furthermore, ML models can be instrumental during large dengue outbreaks, where rapid triage is essential. For instance, an ANN model developed for dengue severity prognosis demonstrated good performance using demographic information and laboratory test results, indicating that ML can quickly predict disease severity and assist in efficient patient triage [20]. Supporting this, other studies have shown high accuracy and effectiveness of ML in dengue diagnosis: an Extra Trees Classifier model achieved over 99% accuracy in Yemen [48], XGBoost reached an AUC of 86% in patients with AFI in Vietnam [49], and Decision Tree and Multilayer Perceptron models attained 98% accuracy in Brazil [50]. Other research on predicting severe COVID-19 in hospitalized children demonstrated higher diagnostic performance with ML approaches compared to LR, with just a few simple and easily collected parameters [22].

The identification of key features such as leukopenia, rash, thrombocytopenia, and age group aligns with clinical knowledge of dengue presentations, validating the models’ ability to capture relevant clinical indicators and suggesting they can effectively complement and enhance existing diagnostic processes. Leukopenia and thrombocytopenia are well-documented hematological manifestations of dengue, indicating the disease’s impact on the blood cell count [51] and were identified as key predictive variables in a similar study [20]. Although these hematological markers substantially contributed to model performance, particularly in smaller feature sets, the XGBoost model with the 48-variable set excluding leukopenia and thrombocytopenia achieved an AUC of 94.1%, demonstrating that strong predictive performance can be attained without relying solely on these laboratory values. This finding highlights the potential for accurate dengue prediction in resource-limited settings where access to CBCs may be restricted. Although IgG testing modestly improved AUCs, its limited availability, potential for misinterpretation in routine clinical practice, and reliance on specialized laboratory infrastructure compared to readily available CBCs restrict its utility as a routine diagnostic tool. The inclusion of age group is consistent with other ML prediction models [20, 22] and highlights the varying clinical presentations and risks across different age demographics, underscoring the need for age-specific diagnostic approaches. Rash, while common in dengue, is also seen in various other febrile illnesses and should be considered alongside other symptoms for differential diagnosis [53]. Similarly, the absence of nasal discharge, more indicative of respiratory pathogens, can also aid in differentiation. Other similar studies also identified body temperature [20] and duration of fever [21] as important features.

Identifying monthly dengue incidence as a key feature underscores the value of temporal patterns for accurate dengue prediction. However, the impact of this data may vary depending on its granularity. While our study highlights potential benefits, other research suggests that using broader regional incidence data may yield smaller improvements compared to more localized data reflecting the specific transmission dynamics in a patient’s area [21]. These findings suggest the need for real-time surveillance and timely intervention strategies, potentially leading to more effective public health responses during peak transmission periods [54]. By leveraging localized monthly incidence data, healthcare systems can better allocate resources, anticipate outbreaks, and implement targeted prevention measures, ultimately reducing the burden of dengue on communities.

Despite the high performance of advanced models like XGBoost, it is noteworthy that simpler models like multivariable LR also achieved AUCs of 91% for the larger feature sets. This indicates that a substantial portion of the predictive power lies within core features, potentially making LR a more interpretable and computationally efficient option for some applications. However, for the larger feature sets, gradient boosting models (XGBoost, LightGBM, AdaBoost) outperformed LR, SVM, and ANN, which may be attributed to the gradient boosting models’ ability to better capture complex feature interactions and non-linear relationships within the data. Gradient boosting models are inherently designed to improve prediction accuracy by iteratively focusing on the hardest-to-predict cases, which can be particularly advantageous when dealing with larger and more complex feature sets. On the other hand, consistently lower AUC values for SVM and ANN across all feature sets in our study suggest potential challenges these models face in capturing intricate feature interactions and effectively generalizing from the data. Specifically, ANNs may struggle with high-dimensional datasets due to the “curse of dimensionality,” where the addition of more features can lead to sparse data, making patterns harder to identify [55]. This, coupled with the risk of overfitting—where the model memorizes the training data but fails to generalize well to unseen data—can reduce accuracy, increase training times, and decrease interpretability. These factors contribute to the observed performance gap between ANNs and the gradient boosting models, particularly as the feature set size grows.

The observation that the performance improvement between the 32-variable and 48-variable sets is minimal suggests a point of diminishing returns. Here, additional features contribute little to predictive power while potentially increasing model complexity and training time. Thus, while ensemble methods like XGBoost may be preferable for applications where maximizing predictive accuracy is paramount, it is important to weigh the benefits of added complexity against the potential gains in performance.

While some models demonstrated high sensitivity, others prioritized specificity, highlighting the need to carefully consider the desired balance between correctly identifying true positives (high sensitivity) and minimizing false positives (high specificity) for specific clinical applications. Another study from Thailand suggests that incorporating dengue NS1 rapid test results enhances diagnostic specificity in models like Bayesian networks, indicating potential for similar improvements in our ML models, which could better confirm non-dengue cases when combined with clinical and laboratory data [21].

This study was subject to several limitations. First, the dataset’s heavy skew towards data from the 2012-2013 outbreak and its primary focus on DENV-1 may limit the generalizability of the models to other periods, regions, or serotypes. Future research should focus on validating these models in diverse settings, integrating real-time data, and exploring the inclusion of newer diagnostic features to refine their accuracy and applicability further. Second, although more features generally improve model performance, there is a risk of overfitting, especially with smaller datasets. Third, the models were developed using data from SEDSS, where the inclusion criteria required febrile illness. This criterion may limit the generalizability of the models to populations without similar inclusion criteria. Fourth, as diagnostic tools, these models would need to be re-fitted to different variables and populations to ensure their accuracy and applicability across various settings. Fifth, the SEDSS dataset used in this study is systematically collected and robust, which may not accurately reflect the conditions of real-world datasets. In many real-world applications, data from electronic health records can be sparse, contain free text fields, or have incomplete information. This could affect the model’s performance if it were trained on less-structured data.

This study demonstrates the potential of ML models, particularly XGBoost and LightGBM, to improve dengue diagnostic accuracy. By incorporating a wider range of features, including temporal patterns like monthly dengue incidence, these models achieved high AUC values, exceeding 90% for larger feature sets. This enables early and precise diagnosis, which could lead to improved patient outcomes and reduced viral spread during outbreaks, particularly in resource-scarce settings where the availability of rapid tests is often constrained. Furthermore, our findings highlight the importance of both complex ensemble methods and simpler models. While XGBoost and LightGBM offer superior performance, even LR achieved strong accuracy with core features. However, careful consideration of model interpretability, generalizability, and continuous validation are crucial for real-world implementation. It is important to note that these ML models are intended to aid and improve dengue diagnosis rather than replace current methods, such as laboratory testing. Future research should focus on addressing these aspects to ensure robust and generalizable ML models that can empower clinicians and public health authorities to effectively manage and control dengue worldwide.

To facilitate clinical application, developing a user-friendly tool such as a calculator, Shiny app, or web-based interface to implement the prediction model is a crucial next step. This would bridge the gap between complex ML models and clinical practice, allowing for efficient integration into routine care and potentially improving diagnostic accuracy and timeliness.

## Data Availability

Data cannot be shared publicly because data cannot be deidentified at the granular level of analyses performed. Data are available from the CDC and PHSU study management team (contact) for
researchers who meet the criteria for access to confidential data.

## Disclaimer

The findings and conclusions in this report are those of the authors and do not necessarily represent the official position of the US Centers for Disease Control and Prevention.

## Notes

### Competing Interest Statement

The authors have declared no competing interest.

### Funding Statement

This research was funded by Centers for Disease Control and Prevention, grant numbers U01CK000473 and U01CK000580 (VRA).

## References

1. Paz-Bailey G, Adams LE, Deen J, Anderson KB, Katzelnick LC. Dengue. Lancet. 2024;403(10427):667–82.

2. Gubler DJ. The global emergence/resurgence of arboviral diseases as public health problems. Archives of medical research. 2002;33(4):330–42.

3. Madewell ZJ. Arboviruses and Their Vectors. South Med J. 2020;113(10):520–3.

4. Simmons CP, Farrar JJ, Nguyen v V, Wills B. Dengue. N Engl J Med. 2012;366(15):1423–32.

5. Toan NT, Rossi S, Prisco G, Nante N, Viviani S. Dengue epidemiology in selected endemic countries: factors influencing expansion factors as estimates of underreporting. Trop Med Int Health. 2015;20(7):840–63.

6. Undurraga EA, Halasa YA, Shepard DS. Use of expansion factors to estimate the burden of dengue in Southeast Asia: a systematic analysis. PLoS Negl Trop Dis. 2013;7(2):e2056.

7. Bhatt S, Gething PW, Brady OJ, Messina JP, Farlow AW, Moyes CL, et al. The global distribution and burden of dengue. Nature. 2013;496(7446):504-7.

8. Global, regional, and national age-sex-specific mortality for 282 causes of death in 195 countries and territories, 1980-2017: a systematic analysis for the Global Burden of Disease Study 2017. Lancet. 2018;392(10159):1736–88.

9. Sharp TM, Ryff KR, Santiago GA, Margolis HS, Waterman SH. Lessons Learned from Dengue Surveillance and Research, Puerto Rico, 1899-2013. Emerg Infect Dis. 2019;25(8):1522–30.

10. Centers for Disease Control and Prevention. ArboNET 2024 [cited 2024 August 19]. Available from: https://www.cdc.gov/mosquitoes/php/arbonet/index.html.

11. Ryff KR, Rivera A, Rodriguez DM, Santiago GA, Medina FA, Ellis EM, et al. Epidemiologic Trends of Dengue in U.S. Territories, 2010-2020. MMWR Surveill Summ. 2023;72(4):1-12.

12. Méndez-Lázaro P, Muller-Karger F, Otis D, Mccarthy M, Peña-Orellana M. Assessing Climate Variability Effects on Dengue Incidence in San Juan, Puerto Rico. International Journal of Environmental Research and Public Health. 2014;11(9):9409–28.

13. Wong JM, Volkman HR, Adams LE, García CO, Martinez-Quiñones A, Perez-Padilla J, et al. Clinical Features of COVID-19, Dengue, and Influenza among Adults Presenting to Emergency Departments and Urgent Care Clinics—Puerto Rico, 2012–2021. The American Journal of Tropical Medicine and Hygiene. 2023;108(1):107.

14. Wong J, Horwitz MM, Zhou L, Toh S. Using machine learning to identify health outcomes from electronic health record data. Curr Epidemiol Rep. 2018;5(4):331–42.

15. Rocha FP, Giesbrecht M. Machine learning algorithms for dengue risk assessment: a case study for São Luís do Maranhão. Computational and Applied Mathematics. 2022;41(8):393.

16. Nasir M, Summerfield NS, Carreiro S, Berlowitz D, Oztekin A. A machine learning approach for diagnostic and prognostic predictions, key risk factors and interactions. Health Services and Outcomes Research Methodology. 2024.

17. Lai CK, Leung E, He Y, Cheung CC, Oliver MOY, Yu Q, et al. A machine learning-based risk score for prediction of infective endocarditis among patients with Staphylococcus aureus bacteraemia - The SABIER score. J Infect Dis. 2024.

18. Tiwari P, Colborn KL, Smith DE, Xing F, Ghosh D, Rosenberg MA. Assessment of a Machine Learning Model Applied to Harmonized Electronic Health Record Data for the Prediction of Incident Atrial Fibrillation. JAMA Netw Open. 2020;3(1):e1919396.

19. Ho TS, Weng TC, Wang JD, Han HC, Cheng HC, Yang CC, et al. Comparing machine learning with case-control models to identify confirmed dengue cases. PLoS Negl Trop Dis. 2020;14(11):e0008843.

20. Huang SW, Tsai HP, Hung SJ, Ko WC, Wang JR. Assessing the risk of dengue severity using demographic information and laboratory test results with machine learning. PLoS Negl Trop Dis. 2020;14(12):e0008960.

21. Sa-Ngamuang C, Haddawy P, Luvira V, Piyaphanee W, Iamsirithaworn S, Lawpoolsri S. Accuracy of dengue clinical diagnosis with and without NS1 antigen rapid test: Comparison between human and Bayesian network model decision. PLoS Negl Trop Dis. 2018;12(6):e0006573.

22. Liu P, Xing Z, Peng X, Zhang M, Shu C, Wang C, et al. Machine learning versus multivariate logistic regression for predicting severe COVID-19 in hospitalized children with Omicron variant infection. J Med Virol. 2024;96(2):e29447.

23. Tomashek KM, Rivera A, Torres-Velasquez B, Hunsperger EA, Munoz-Jordan JL, Sharp TM, et al. Enhanced Surveillance for Fatal Dengue-Like Acute Febrile Illness in Puerto Rico, 2010-2012. PLoS Negl Trop Dis. 2016;10(10):e0005025.

24. Madewell ZJ, Hernandez-Romieu AC, Wong JM, Zambrano LD, Volkman HR, Perez-Padilla J, et al. Sentinel Enhanced Dengue Surveillance System - Puerto Rico, 2012-2022. MMWR Surveill Summ. 2024;73(3):1–29.

25. Read JS, Torres-Velasquez B, Lorenzi O, Rivera Sanchez A, Torres-Torres S, Rivera LV, et al. Symptomatic Zika Virus Infection in Infants, Children, and Adolescents Living in Puerto Rico. JAMA Pediatr. 2018;172(7):686–93.

26. Harris PA, Taylor R, Minor BL, Elliott V, Fernandez M, O’Neal L, et al. The REDCap consortium: Building an international community of software platform partners. J Biomed Inform. 2019;95:103208.

27. Harris PA, Taylor R, Thielke R, Payne J, Gonzalez N, Conde JG. Research electronic data capture (REDCap)--a metadata-driven methodology and workflow process for providing translational research informatics support. J Biomed Inform. 2009;42(2):377–81.

28. Siriseriwan W. smotefamily: A Collection of Oversampling Techniques for Class Imbalance Problem Based on SMOTE 2024 [cited 2024 June 17]. Available from: https://cran.r-project.org/web/packages/smotefamily/index.html.

29. Kuhn M. caret: Classification and Regression Training 2023 [cited 2024 June 17]. Available from: https://cran.r-project.org/web/packages/caret/index.html.

30. Breiman L. Random forests. Machine learning. 2001;45:5–32.

31. Hearst MA, Dumais ST, Osuna E, Platt J, Scholkopf B. Support vector machines. IEEE Intelligent Systems and their applications. 1998;13(4):18–28.

32. Yegnanarayana B. Artificial neural networks: PHI Learning Pvt. Ltd.; 2009.

33. Schapire RE. The boosting approach to machine learning: An overview. Nonlinear estimation and classification. 2003:149–71.

34. Bentéjac C, Csörgő A, Martínez-Muñoz G. A comparative analysis of gradient boosting algorithms. Artificial Intelligence Review. 2021;54:1937–67.

35. Breiman L. randomForest: Breiman and Cutler’s Random Forests for Classification and Regression 2022 [cited 2024 June 17]. Available from: https://cran.r-project.org/web/packages/randomForest/index.html.

36. Meyer D. e1071: Misc Functions of the Department of Statistics, Probability Theory Group (Formerly: E1071), TU Wien 2023 [cited 2024 June 20]. Available from: https://cran.r-project.org/web/packages/e1071/index.html.

37. Ripley B. nnet: Feed-Forward Neural Networks and Multinomial Log-Linear Models 2023 [cited 2024 June 18]. Available from: https://cran.r-project.org/web/packages/nnet/index.html.

38. Culp M. ada: The R Package Ada for Stochastic Boosting 2016 [cited 2024 June 18]. Available from: https://cran.r-project.org/web/packages/ada/index.html.

39. Shi Y. lightgbm: Light Gradient Boosting Machine 2024 [cited 2024 June 17]. Available from: https://cran.r-project.org/web/packages/lightgbm/index.html.

40. Chen T. xgboost: Extreme Gradient Boosting 2024 [cited 2024 June 17]. Available from: https://cran.r-project.org/web/packages/xgboost/index.html.

41. DeLong ER, DeLong DM, Clarke-Pearson DL. Comparing the areas under two or more correlated receiver operating characteristic curves: a nonparametric approach. Biometrics. 1988;44(3):837–45.

42. Youden WJ. Index for rating diagnostic tests. Cancer. 1950;3(1):32–5.

43. Altmann A, Toloşi L, Sander O, Lengauer T. Permutation importance: a corrected feature importance measure. Bioinformatics. 2010;26(10):1340–7.

44. R Core Team. R: A language and environment for statistical computing. Vienna, Austria: R Foundation for Statistical Computing; 2024 [

45. Raafat N, Loganathan S, Mukaka M, Blacksell SD, Maude RJ. Diagnostic accuracy of the WHO clinical definitions for dengue and implications for surveillance: A systematic review and meta-analysis. PLoS Negl Trop Dis. 2021;15(4):e0009359.

46. Sigera PC, Amarasekara R, Rodrigo C, Rajapakse S, Weeratunga P, De Silva NL, et al. Risk prediction for severe disease and better diagnostic accuracy in early dengue infection; the Colombo dengue study. BMC Infect Dis. 2019;19(1):680.

47. Mejía MFÁ, Shu P-Y, Ji D-D. Accuracy of Dengue, Chikungunya, and Zika diagnoses by primary healthcare physicians in Tegucigalpa, Honduras. BMC Infectious Diseases. 2023;23(1):371.

48. Abdualgalil B, Abraham S, M. Ismael W. Early Diagnosis for Dengue Disease Prediction Using Efficient Machine Learning Techniques Based on Clinical Data. 2022. 2022;3(3):12.

49. Ming DK, Tuan NM, Hernandez B, Sangkaew S, Vuong NL, Chanh HQ, et al. The Diagnosis of Dengue in Patients Presenting With Acute Febrile Illness Using Supervised Machine Learning and Impact of Seasonality. Front Digit Health. 2022;4:849641.

50. Bohm BC, Borges FEM, Silva SCM, Soares AT, Ferreira DD, Belo VS, et al. Utilization of machine learning for dengue case screening. BMC Public Health. 2024;24(1):1573.

51. Vogt MB, Lahon A, Arya RP, Spencer Clinton JL, Rico-Hesse R. Dengue viruses infect human megakaryocytes, with probable clinical consequences. PLoS Negl Trop Dis. 2019;13(11):e0007837.

52. Teo A, Tan HD, Loy T, Chia PY, Chua CLL. Understanding antibody-dependent enhancement in dengue: Are afucosylated IgG1s a concern? PLoS Pathog. 2023;19(3):e1011223.

53. Daumas RP, Passos SRL, Oliveira RVC, Nogueira RMR, Georg I, Marzochi KBF, Brasil P. Clinical and laboratory features that discriminate dengue from other febrile illnesses: a diagnostic accuracy study in Rio de Janeiro, Brazil. BMC Infectious Diseases. 2013;13(1):77.

54. Madewell ZJ, Hemme RR, Adams L, Barrera R, Waterman SH, Johansson MA. Comparing vector and human surveillance strategies to detect arbovirus transmission: A simulation study for Zika virus detection in Puerto Rico. PLoS Negl Trop Dis. 2019;13(12):e0007988.

55. Grohs P, Ibragimov S, Jentzen A, Koppensteiner S. Lower bounds for artificial neural network approximations: A proof that shallow neural networks fail to overcome the curse of dimensionality. Journal of Complexity. 2023;77:101746.

