## Supplement for "Machine learning for improved dengue diagnosis, Puerto Rico"

**S1 Table.** Odds of dengue infection for selected characteristics, SEDSS, May 2012–June 2024 (N = 49679).

**S2 Table**. Machine learning models and hyperparameter tuning.

**S3 Table.** Additional pathogens in study population, SEDSS, May 2012–June 2024 (N = 49679).

**S4 Table.** Performance of each algorithm for predicting dengue infection on the training set, SEDSS, May 2012–June 2024.

**S5 Table.** Performance of eXtreme Gradient Boosting Model for predicting dengue infection on the test set 1) with IgG testing and 2) without leukopenia and thrombocytopenia, SEDSS, May 2012–June 2024.

**S6 Table.** Adjusted odds of dengue infection for characteristics identified by stepwise selection from the 48-variable feature set, SEDSS, May 2012–June 2024.

**S1 Fig.** Feature importance for 8-variable feature set for each machine learning model, SEDSS, May 2012–June 2024.

**S2 Fig.** Feature importance for 20-variable feature set for each machine learning model, SEDSS, May 2012–June 2024.

**S3 Fig.** Feature importance for 32-variable feature set for each machine learning model, SEDSS, May 2012–June 2024.

**S4 Fig.** Feature importance for 48-variable feature set for each machine learning model, SEDSS, May 2012–June 2024.

**S5 Fig**. Incremental improvement in AUC by sequentially adding features identified from XGBoost models, SEDSS, May 2012–June 2024.

| **S1 Table.** Crude odds of dengue infection compared to non-dengue cases for selected characteristics, SEDSS, May 2012–June 2024 (N = 49679). | | |
| --- | --- | --- |
|  | **OR (95% CI)** | **p-value** |
| **Month (ref: January)** |  | <0.001 |
| February | 0.74 (0.57, 0.96) |  |
| March | 0.50 (0.37, 0.66) |  |
| April | 0.54 (0.41, 0.72) |  |
| May | 0.69 (0.53, 0.89) |  |
| June | 1.00 (0.79, 1.27) |  |
| July | 0.93 (0.73, 1.18) |  |
| August | 1.06 (0.85, 1.34) |  |
| September | 1.04 (0.83, 1.32) |  |
| October | 1.02 (0.81, 1.29) |  |
| November | 1.34 (1.07, 1.68) |  |
| December | 1.10 (0.87, 1.40) |  |
| **Days post onset (ref: 0)** |  | <0.001 |
| 1-3 | 1.70 (1.41, 2.07) |  |
| 4-6 | 7.19 (5.95, 8.76) |  |
| 7+ | 2.61 (1.92, 3.51) |  |
| **Age group (ref: <1)** |  | <0.001 |
| 1-4 | 1.13 (0.77, 1.72) |  |
| 5-9 | 4.03 (2.81, 5.98) |  |
| 10-19 | 10.90 (7.72, 15.99) |  |
| 20-29 | 4.10 (2.85, 6.11) |  |
| 30-39 | 3.05 (2.07, 4.64) |  |
| 40-49 | 3.03 (2.03, 4.64) |  |
| 50+ | 2.61 (1.81, 3.89) |  |
| **Male sex (ref: female)** | 1.30 (1.18, 1.44) | <0.001 |
| **Health region (ref: Ponce)** |  | <0.001 |
| Aguadilla | 1.56 (0.25, 5.07) |  |
| Arecibo | 0.68 (0.21, 1.60) |  |
| Bayamon | 0.84 (0.55, 1.22) |  |
| Caguas | 1.64 (1.09, 2.36) |  |
| Fajardo | 1.53 (0.89, 2.44) |  |
| Mayaguez | 1.75 (0.69, 3.66) |  |
| Metro | 1.87 (1.65, 2.11) |  |
| Unknown | 0.23 (0.01, 1.04) |  |
| **Traveled to another country in past two weeks (ref: no)** | 1.54 (1.21, 1.95) | <0.001 |
| **Household member with dengue (ref: no)** | 8.64 (7.06, 10.50) | <0.001 |
| **Reported mosquito bites in past month** | 3.11 (2.80, 3.44) | <0.001 |
| **Comorbidities** |  |  |
| Chronic pulmonary disease or asthma | 1.04 (0.92, 1.18) | 0.525 |
| Cancer | 0.86 (0.57, 1.25) | 0.450 |
| Chronic kidney disease | 0.59 (0.31, 0.99) | 0.048 |
| Congenital heart disease | 0.86 (0.64, 1.12) | 0.266 |
| Diabetes | 0.76 (0.62, 0.93) | 0.006 |
| High cholesterol | 0.78 (0.62, 0.98) | 0.029 |
| Hypertension | 0.63 (0.53, 0.74) | <0.001 |
| Arthritis | 0.32 (0.21, 0.48) | <0.001 |
| Thyroid disease | 0.68 (0.53, 0.86) | 0.001 |
| Obesity | 0.60 (0.51, 0.69) | <0.001 |
| Gastritis | 1.58 (1.04, 2.28) | 0.032 |
| **Warning signs** |  |  |
| Persistent vomiting | 1.44 (1.29, 1.62) | <0.001 |
| Abdominal pain | 2.78 (2.52, 3.07) | <0.001 |
| Restlessness | 1.50 (1.36, 1.66) | <0.001 |
| Clinical fluid accumulation | 0.89 (0.71, 1.10) | 0.278 |
| Mucosal bleeding | 2.02 (1.75, 2.31) | <0.001 |
| Hepatomegaly | 17.63 (12.03, 25.54) | <0.001 |
| **Other clinical signs** |  |  |
| Fever | 19.10 (10.56, 39.83) | <0.001 |
| Conjunctivitis | 2.51 (2.08, 3.01) | <0.001 |
| Chills | 3.53 (3.13, 3.99) | <0.001 |
| Nausea | 2.60 (2.34, 2.89) | <0.001 |
| No appetite | 2.42 (2.15, 2.72) | <0.001 |
| Rash | 6.14 (5.55, 6.79) | <0.001 |
| Yellow skin | 2.82 (2.17, 3.61) | <0.001 |
| Itchy skin | 4.10 (3.69, 4.54) | <0.001 |
| Headache | 3.72 (3.26, 4.27) | <0.001 |
| Eye pain | 4.00 (3.62, 4.43) | <0.001 |
| Myalgia | 3.24 (2.89, 3.62) | <0.001 |
| Arthralgia | 2.53 (2.29, 2.80) | <0.001 |
| Tachypnea | 3.50 (3.16, 3.86) | <0.001 |
| Back pain | 2.12 (1.92, 2.34) | <0.001 |
| Calf pain | 1.75 (1.58, 1.94) | <0.001 |
| Arthritis | 1.95 (1.69, 2.24) | <0.001 |
| Nasal discharge | 0.29 (0.26, 0.32) | <0.001 |
| Sore throat | 0.68 (0.61, 0.75) | <0.001 |
| Cough | 0.34 (0.30, 0.37) | <0.001 |
| Diarrhea | 2.03 (1.84, 2.25) | <0.001 |
| Hypotension | 3.46 (2.86, 4.16) | <0.001 |
| Narrow pulse pressure | 1.46 (1.04, 1.99) | 0.031 |
| Seizure | 1.32 (0.88, 1.89) | 0.176 |
| Capillary refill <3 | 1.02 (0.82, 1.24) | 0.878 |
| Pale skin | 3.18 (2.88, 3.51) | <0.001 |
| Blue lips | 3.18 (2.45, 4.06) | <0.001 |
| **Laboratory** |  |  |
| Leukopenia | 23.37 (21.03, 25.98) | <0.001 |
| Thrombocytopenia | 8.83 (7.97, 9.78) | <0.001 |
| **Monthly dengue incidence per 100,000 people (10-unit increase)** | 2.23 (2.17, 2.30) | <0.001 |

| **S2 Table. Machine learning models and hyperparameter tuning.** To optimize the performance of various machine learning models for predicting a dengue diagnosis, a grid search strategy was employed across several algorithms. The details of the grid search strategy and the specific hyperparameters evaluated for each model are outlined below. We also included the optimal values identified through this process that were included in the final models. | | | | | | |
| --- | --- | --- | --- | --- | --- | --- |
| Model, R package | Parameter | Definition | Included in final model  8-variable set | Included in final model  20-variable set | Included in final model  32-variable set | Included in final model  48-variable set |
| XGBoost,  xgboost [40] | booster type | specifies type of model to use | decision tree | decision tree | decision tree | decision tree |
|  | objective | defines the learning task and the corresponding loss function | binary:logistic | binary:logistic | binary:logistic | binary:logistic |
|  | nrounds | number of boosting iterations | 1000 | 1000 | 1000 | 1000 |
|  | max_depth | maximum depth of a tree | 2 | 2 | 3 | 7 |
|  | eta | learning rate | 0.20 | 0.07 | 0.25 | 0.19 |
|  | gamma | minimum loss reduction required to make a further partition on a leaf node of the tree | 0 | 0 | 0 | 0 |
|  | colsample_bytree | subsample ratio of columns when constructing each tree | 0.7 | 0.5 | 1 | 0.4 |
|  | min_child_weight | minimum sum of instance weight (hessian) needed in a child | 1 | 2 | 1.1 | 1 |
| LightGBM,  lightgbm [39] | booster type | specifies type of model to use | decision tree | decision tree | decision tree | decision tree |
|  | objective | defines the learning task | binary | binary | Binary | binary |
|  | num_leaves | maximum tree leaves for base learners | 30 | 60 | 65 | 30 |
|  | max_depth | maximum depth of a tree | -1 (no limit) | -1 (no limit) | -1 (no limit) | -1 (no limit) |
|  | learning_rate | shrinks the contribution of each tree by learning_rate | 0.1 | 0.1 | 0.1 | 0.1 |
|  | nrounds | number of boosting iterations | 1000 | 1000 | 1000 | 1000 |
|  | min_split_gain | minimum gain to make a split | 0 | 0 | 0 | 0 |
|  | feature_fraction | randomly select a subset of features for each tree during training | 0.70 | 0.45 | 0.80 | 0.80 |
|  | bagging_fraction | randomly sample fraction of data used for each training iteration | 0.90 | 0.96 | 0.80 | 0.80 |
|  | bagging_freq | perform bagging at every k iteration | 6 | 5 | 6 | 5 |
| AdaBoost, ada [38] | type | type of boosting algorithm | discrete boosting | discrete boosting | discrete boosting | discrete boosting |
|  | nu | shrinkage parameter for boosting | 0.1 | 0.1 | 0.1 | 0.1 |
|  | iter | number of boosting iterations | 100 | 100 | 100 | 100 |
|  | loss | loss function used to evaluate the model’s performance | exponential | exponential | exponential | exponential |
| SVM, e1071 [36] | kernel type | specifies kernel function used to map data into a higher-dimensional space | linear | linear | linear | linear |
|  | Gamma | Kernel coefficient | 1/n_features = 0.125 | 1/n_features = 0.050 | 1/n_features = 0.031 | 1/n_features = 0.021 |
|  | Cost | controls trade-off between maximizing the margin and minimizing classification errors | 1 | 1 | 1 | 1 |
|  | probability | enables probability predictions | TRUE | TRUE | TRUE | TRUE |
| ANN, nnet [37] | size | number of units in the hidden layer | 10 | 10 | 10 | 10 |
|  | decay | weight decay for regularization to avoid overfitting | 0.10 | 0.40 | 0.45 | 0.55 |
|  | rang | initial range of weights | 0.5 | 0.6 | 0.6 | 0.6 |
|  | maxit | maximum number of iterations | 500 | 500 | 500 | 500 |
|  | entropy | maximum conditional likelihood | least-squares error | least-squares error | least-squares error | least-squares error |
| RF, randomForest [35] | ntree | number of trees in forest | 500 | 500 | 500 | 500 |
|  | mtry | controls randomness of forest | 2 | 2 | 26 | 38 |

| **S3 Table.** Additional pathogens in study population, SEDSS, May 2012–June 2024 (N = 49679**)**. | |
| --- | --- |
|  | **N (%)** |
| **Tested for chikungunya virus** |  |
| Positive | 2291 ( 8.4) |
| Negative | 24901 (91.6) |
| **Tested for Zika virus** |  |
| Positive | 1899 ( 8.8) |
| Negative | 19651 (91.2) |
| **Tested for influenza A virus** |  |
| Positive | 4296 ( 9.4) |
| Negative | 41555 (90.6) |
| **Tested for influenza B virus** |  |
| Positive | 1726 ( 3.8) |
| Negative | 44126 (96.2) |
| **Tested for respiratory syncytial virus** |  |
| Positive | 1576 ( 3.4) |
| Negative | 44402 (96.6) |
| **Tested for human metapneumovirus** |  |
| Positive | 1043 ( 2.3) |
| Negative | 44936 (97.7) |
| **Tested for human adenovirus** |  |
| Positive | 1835 ( 4.0) |
| Negative | 44144 (96.0) |
| **Tested for human parainfluenza virus 1** |  |
| Positive | 478 ( 1.0) |
| Negative | 45498 (99.0) |
| **Tested for human coronavirus** |  |
| Positive | 46 ( 2.5) |
| Negative | 1786 (97.5) |
| **Tested for SARS-CoV-2** |  |
| Positive | 2322 (15.2) |
| Negative | 12968 (84.8) |

| **S4 Table.** Performance of each algorithm for predicting dengue infection on the **training set**, SEDSS, May 2012–June 2024. | | | | | | | | |
| --- | --- | --- | --- | --- | --- | --- | --- | --- |
| **Model** | **Accuracy** | **Sensitivity** | **Specificity** | **PPV** | **NPV** | **F1 Score** | **Kappa** | **AUC** |
| **8-variable feature set** |  |  |  |  |  |  |  |  |
| Logistic Regression | 0.8188 | 0.9085 | 0.7291 | 0.7703 | 0.8885 | 0.8337 | 0.6376 | 0.8825 |
| Support Vector Machine | 0.7853 | 0.8693 | 0.7012 | 0.7442 | 0.8429 | 0.8019 | 0.5706 | 0.8697 |
| Random Forest | 0.8158 | 0.9024 | 0.7291 | 0.7691 | 0.8820 | 0.8305 | 0.6315 | 0.8789 |
| Artificial Neural Network | 0.8240 | 0.9111 | 0.7369 | 0.7760 | 0.8924 | 0.8381 | 0.6481 | 0.8899 |
| Adaptive Boosting | 0.8179 | 0.8763 | 0.7596 | 0.7847 | 0.8600 | 0.8280 | 0.6359 | 0.8909 |
| Light Gradient Boosting | 0.8249 | 0.9190 | 0.7308 | 0.7735 | 0.9002 | 0.8400 | 0.6498 | 0.8929 |
| eXtreme Gradient Boosting | 0.8206 | 0.9138 | 0.7274 | 0.7702 | 0.8940 | 0.8359 | 0.6411 | 0.8871 |
| **20-variable feature set** |  |  |  |  |  |  |  |  |
| Logistic Regression | 0.8280 | 0.8737 | 0.7822 | 0.8005 | 0.8610 | 0.8355 | 0.6559 | 0.9067 |
| Support Vector Machine | 0.8210 | 0.8763 | 0.7657 | 0.7890 | 0.8609 | 0.8304 | 0.6420 | 0.8794 |
| Random Forest | 0.8645 | 0.9242 | 0.8049 | 0.8257 | 0.9139 | 0.8721 | 0.7291 | 0.9341 |
| Artificial Neural Network | 0.8415 | 0.8632 | 0.8197 | 0.8272 | 0.8570 | 0.8448 | 0.6829 | 0.9119 |
| Adaptive Boosting | 0.8389 | 0.8659 | 0.8118 | 0.8215 | 0.8582 | 0.8431 | 0.6777 | 0.9286 |
| Light Gradient Boosting | 0.8567 | 0.8789 | 0.8345 | 0.8415 | 0.8733 | 0.8598 | 0.7134 | 0.9310 |
| eXtreme Gradient Boosting | 0.8332 | 0.8345 | 0.8319 | 0.8323 | 0.8341 | 0.8334 | 0.6664 | 0.9132 |
| **32-variable feature set** |  |  |  |  |  |  |  |  |
| Logistic Regression | 0.8637 | 0.8693 | 0.8580 | 0.8596 | 0.8678 | 0.8644 | 0.7274 | 0.9350 |
| Support Vector Machine | 0.8628 | 0.8981 | 0.8275 | 0.8389 | 0.8903 | 0.8675 | 0.7256 | 0.9329 |
| Random Forest | 1 | 1 | 1 | 1 | 1 | 1 | 1 | 1 |
| Artificial Neural Network | 0.8916 | 0.9129 | 0.8702 | 0.8755 | 0.909 | 0.8938 | 0.7831 | 0.9491 |
| Adaptive Boosting | 0.9294 | 0.9382 | 0.9207 | 0.9221 | 0.9371 | 0.9301 | 0.8589 | 0.9822 |
| Light Gradient Boosting | 0.8567 | 0.8789 | 0.8345 | 0.8415 | 0.8733 | 0.8598 | 0.7134 | 0.9947 |
| eXtreme Gradient Boosting | 0.9338 | 0.9547 | 0.9129 | 0.9164 | 0.9527 | 0.9352 | 0.8676 | 0.9840 |
| **48-variable feature set** |  |  |  |  |  |  |  |  |
| Logistic Regression | 0.8676 | 0.8406 | 0.8946 | 0.8886 | 0.8488 | 0.8639 | 0.7352 | 0.9393 |
| Support Vector Machine | 0.8693 | 0.9068 | 0.8319 | 0.8436 | 0.8992 | 0.8741 | 0.7387 | 0.9376 |
| Random Forest | 1 | 1 | 1 | 1 | 1 | 1 | 1 | 1 |
| Artificial Neural Network | 0.9029 | 0.9181 | 0.8876 | 0.8910 | 0.9155 | 0.9043 | 0.8057 | 0.9552 |
| Adaptive Boosting | 0.9456 | 0.9521 | 0.9390 | 0.9398 | 0.9515 | 0.9459 | 0.8911 | 0.9840 |
| Light Gradient Boosting | 0.9608 | 0.9652 | 0.9564 | 0.9568 | 0.9649 | 0.9610 | 0.9216 | 0.9930 |
| eXtreme Gradient Boosting | 0.9647 | 0.9747 | 0.9547 | 0.9556 | 0.9742 | 0.9651 | 0.9294 | 0.9941 |
| **PPV:** positive predictive value; **NPV**: negative predictive value; **AUC**: area under receiver operating characteristic curve. | | | | | | | | |

| **S5 Table.** Performance of eXtreme Gradient Boosting Model for predicting dengue infection on the test set 1) with IgG testing and 2) without leukopenia and thrombocytopenia, SEDSS, May 2012–June 2024. | | | | | | | | |
| --- | --- | --- | --- | --- | --- | --- | --- | --- |
| **Model** | **Accuracy** | **Sensitivity** | **Specificity** | **PPV** | **NPV** | **F1 Score** | **Kappa** | **AUC** |
| **With IgG testing** |  |  |  |  |  |  |  |  |
| 8-variable feature set | 0.8374 | 0.8862 | 0.7886 | 0.8074 | 0.8739 | 0.8450 | 0.6748 | 0.8972 |
| 20-variable feature set | 0.8435 | 0.9024 | 0.7846 | 0.8073 | 0.8894 | 0.8522 | 0.6870 | 0.9145 |
| 32-variable feature set | 0.8882 | 0.9228 | 0.8537 | 0.8631 | 0.9170 | 0.8919 | 0.7764 | 0.9472 |
| 48-variable feature set | 0.8872 | 0.9472 | 0.8272 | 0.8457 | 0.9400 | 0.8936 | 0.7744 | 0.9493 |
| **Without leukopenia and thrombocytopenia** |  |  |  |  |  |  |  |  |
| 8-variable feature set | 0.7439 | 0.7480 | 0.7398 | 0.7419 | 0.7459 | 0.7449 | 0.4878 | 0.8158 |
| 20-variable feature set | 0.8018 | 0.7764 | 0.8272 | 0.8180 | 0.7872 | 0.7967 | 0.6037 | 0.8675 |
| 32-variable feature set | 0.8699 | 0.8780 | 0.8618 | 0.8640 | 0.8760 | 0.8710 | 0.7398 | 0.9353 |
| 48-variable feature set | 0.8811 | 0.8557 | 0.9065 | 0.9015 | 0.8627 | 0.8780 | 0.7622 | 0.9408 |

| **S6 Table.** Adjusted odds of dengue infection for characteristics identified by stepwise selection from the 48-variable feature set, SEDSS, May 2012–June 2024 (N = 49679). | | |
| --- | --- | --- |
|  | **aOR (95% CI)** | **p-value** |
| **Days post onset (ref: 0)** |  | <0.001 |
| 1-3 | 1.32 (0.90, 1.96) |  |
| 4-6 | 2.88 (1.81, 4.62) |  |
| 7+ | 4.10 (1.86, 9.10) |  |
| **Age group (ref: <1)** |  | <0.001 |
| 1-4 | 1.48 (0.68, 3.40) |  |
| 5-9 | 2.49 (1.12, 5.79) |  |
| 10-19 | 4.50 (2.03, 10.43) |  |
| 20-29 | 2.58 (1.09, 6.32) |  |
| 30-39 | 2.17 (0.86, 5.65) |  |
| 40-49 | 2.65 (1.02, 7.04) |  |
| 50+ | 2.68 (1.16, 6.44) |  |
| **Household member with dengue (ref: no)** | 5.43 (2.28, 13.98) | <0.001 |
| **Reported mosquito bites in past month** | 2.01 (1.45, 2.81) | <0.001 |
| **Warning signs** |  |  |
| Persistent vomiting | 0.55 (0.38, 0.81) | <0.001 |
| Restlessness | 0.67 (0.49, 0.90) | <0.001 |
| **Other clinical signs** |  |  |
| Fever | 5.03 (2.04, 14.93) | <0.001 |
| Conjunctivitis | 1.96 (1.11, 3.47) | <0.001 |
| Chills | 1.43 (1.01, 2.01) | <0.001 |
| Nausea | 1.29 (0.94, 1.77) | <0.001 |
| No appetite | 1.41 (1.03, 1.94) | <0.001 |
| Rash | 2.36 (1.70, 3.30) | <0.001 |
| Itchy skin | 1.56 (1.06, 2.30) | <0.001 |
| Eye pain | 1.49 (1.09, 2.04) | <0.001 |
| Myalgia | 1.36 (0.91, 2.04) | <0.001 |
| Arthralgia | 1.34 (0.89, 2.01) | <0.001 |
| Tachypnea | 1.37 (1.00, 1.89) | <0.001 |
| Back pain | 0.65 (0.44, 0.96) | <0.001 |
| Nasal discharge | 0.68 (0.50, 0.92) | <0.001 |
| Sore throat | 0.53 (0.39, 0.72) | <0.001 |
| Cough | 0.68 (0.50, 0.93) | <0.001 |
| Pale skin | 1.32 (0.96, 1.81) | <0.001 |
| **Laboratory** |  |  |
| Leukopenia | 4.93 (3.49, 7.01) | <0.001 |
| Thrombocytopenia | 2.36 (1.57, 3.58) | <0.001 |
| **Monthly dengue incidence per 100,000 people (10-unit increase)** | 2.63 (2.28, 3.05) | <0.001 |

**S1 Fig. Feature importance for 8-variable feature set for each machine learning model, SEDSS, May 2012–June 2024**. The feature importance metrics differ across the various machine learning models depicted. In Random Forest, AdaBoost, LightGBM, and XGBoost, feature importance is measured as the mean decrease in impurity or gain, indicating each feature’s contribution to reducing the overall prediction error. For Support Vector Machine, feature importance is derived from the magnitude of the coefficients of the support vectors, reflecting the influence of features on the decision boundary. Artificial Neural Network uses permutation importance, which quantifies the impact of feature perturbation on model performance. Consequently, these importance scores are on different scales and represent distinct notions of feature contribution, and therefore, direct comparisons across models should be made cautiously.

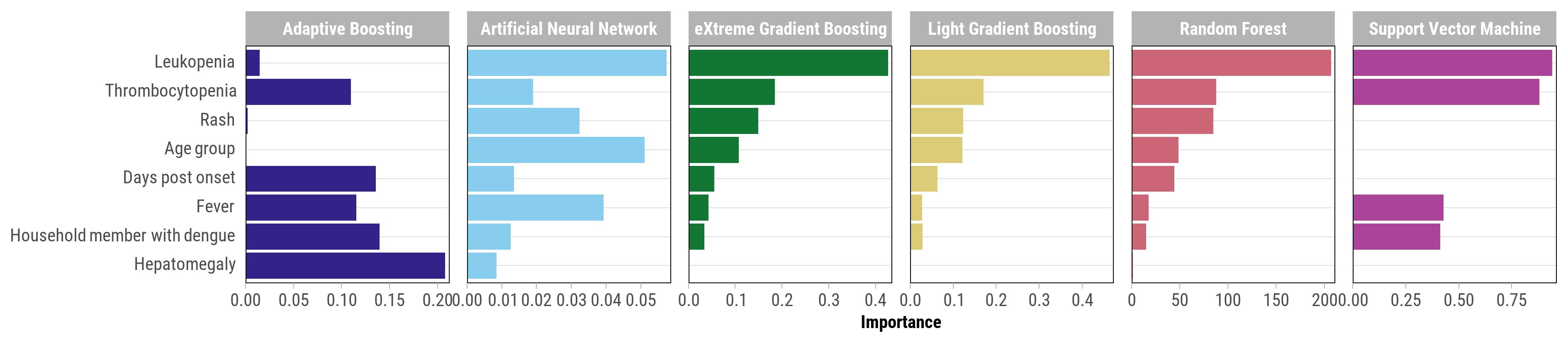

**S2 Fig. Feature importance for 20-variable feature set for each machine learning model, SEDSS, May 2012–June 2024**. The feature importance metrics differ across the various machine learning models depicted. In Random Forest, AdaBoost, LightGBM, and XGBoost, feature importance is measured as the mean decrease in impurity or gain, indicating each feature’s contribution to reducing the overall prediction error. For Support Vector Machine, feature importance is derived from the magnitude of the coefficients of the support vectors, reflecting the influence of features on the decision boundary. Artificial Neural Network uses permutation importance, which quantifies the impact of feature perturbation on model performance. Consequently, these importance scores are on different scales and represent distinct notions of feature contribution, and therefore, direct comparisons across models should be made cautiously.

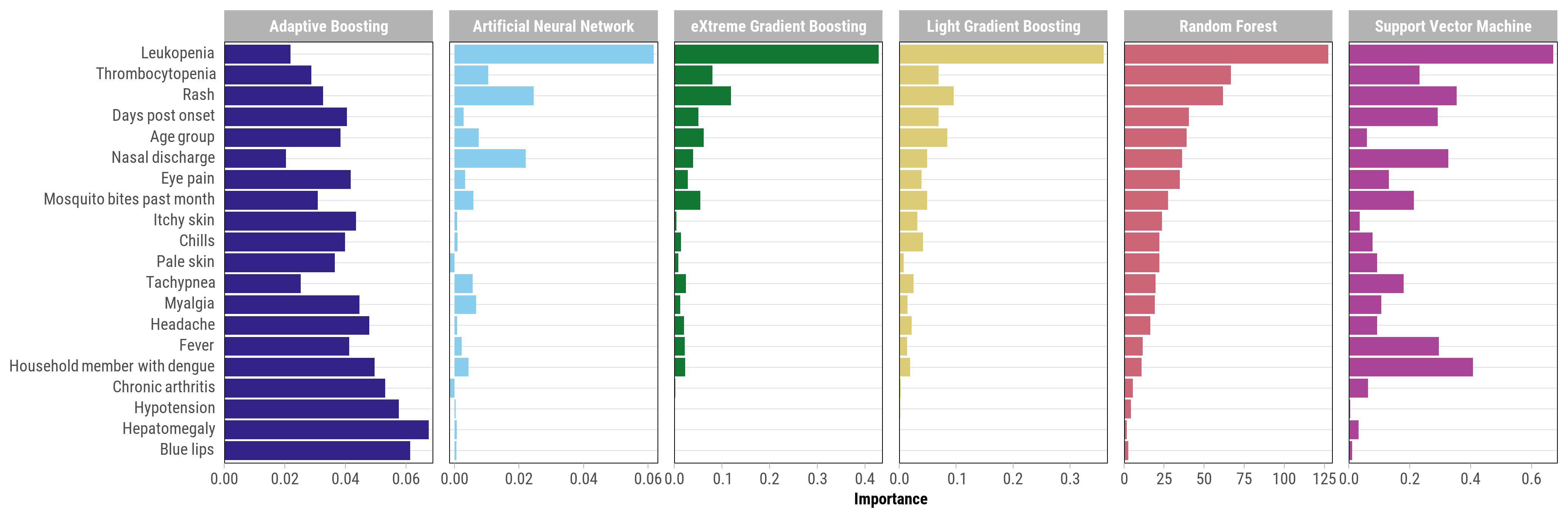

**S3 Fig. Feature importance for 32-variable feature set for each machine learning model, SEDSS, May 2012–June 2024**. The feature importance metrics differ across the various machine learning models depicted. In Random Forest, AdaBoost, LightGBM, and XGBoost, feature importance is measured as the mean decrease in impurity or gain, indicating each feature’s contribution to reducing the overall prediction error. For Support Vector Machine, feature importance is derived from the magnitude of the coefficients of the support vectors, reflecting the influence of features on the decision boundary. Artificial Neural Network uses permutation importance, which quantifies the impact of feature perturbation on model performance. Consequently, these importance scores are on different scales and represent distinct notions of feature contribution, and therefore, direct comparisons across models should be made cautiously.

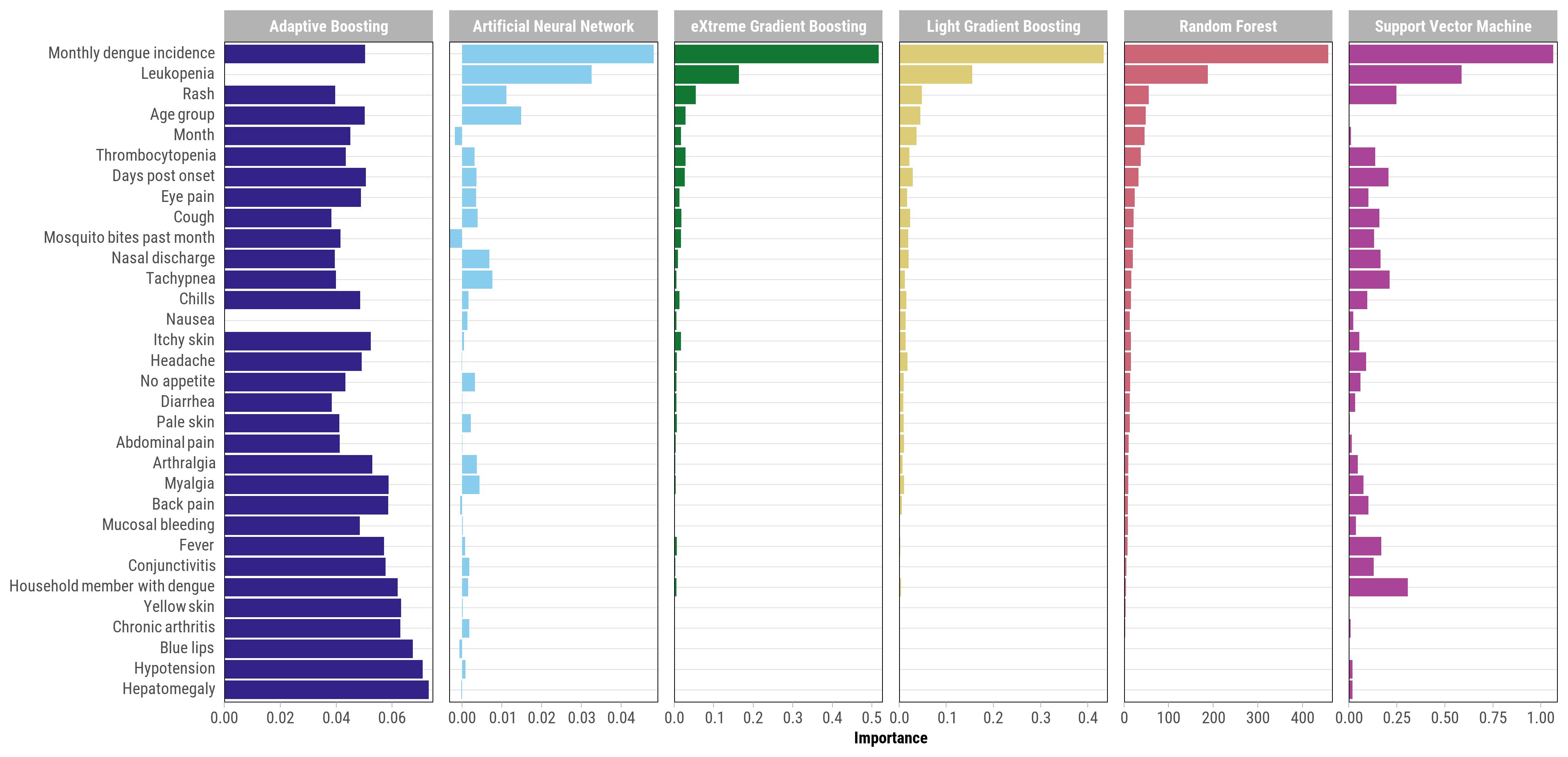

**S4 Fig. Feature importance for 48-variable feature set for each machine learning model, SEDSS, May 2012–June 2024**. The feature importance metrics differ across the various machine learning models depicted. In Random Forest, AdaBoost, LightGBM, and XGBoost, feature importance is measured as the mean decrease in impurity or gain, indicating each feature’s contribution to reducing the overall prediction error. For Support Vector Machine, feature importance is derived from the magnitude of the coefficients of the support vectors, reflecting the influence of features on the decision boundary. Artificial Neural Network uses permutation importance, which quantifies the impact of feature perturbation on model performance. Consequently, these importance scores are on different scales and represent distinct notions of feature contribution, and therefore, direct comparisons across models should be made cautiously.

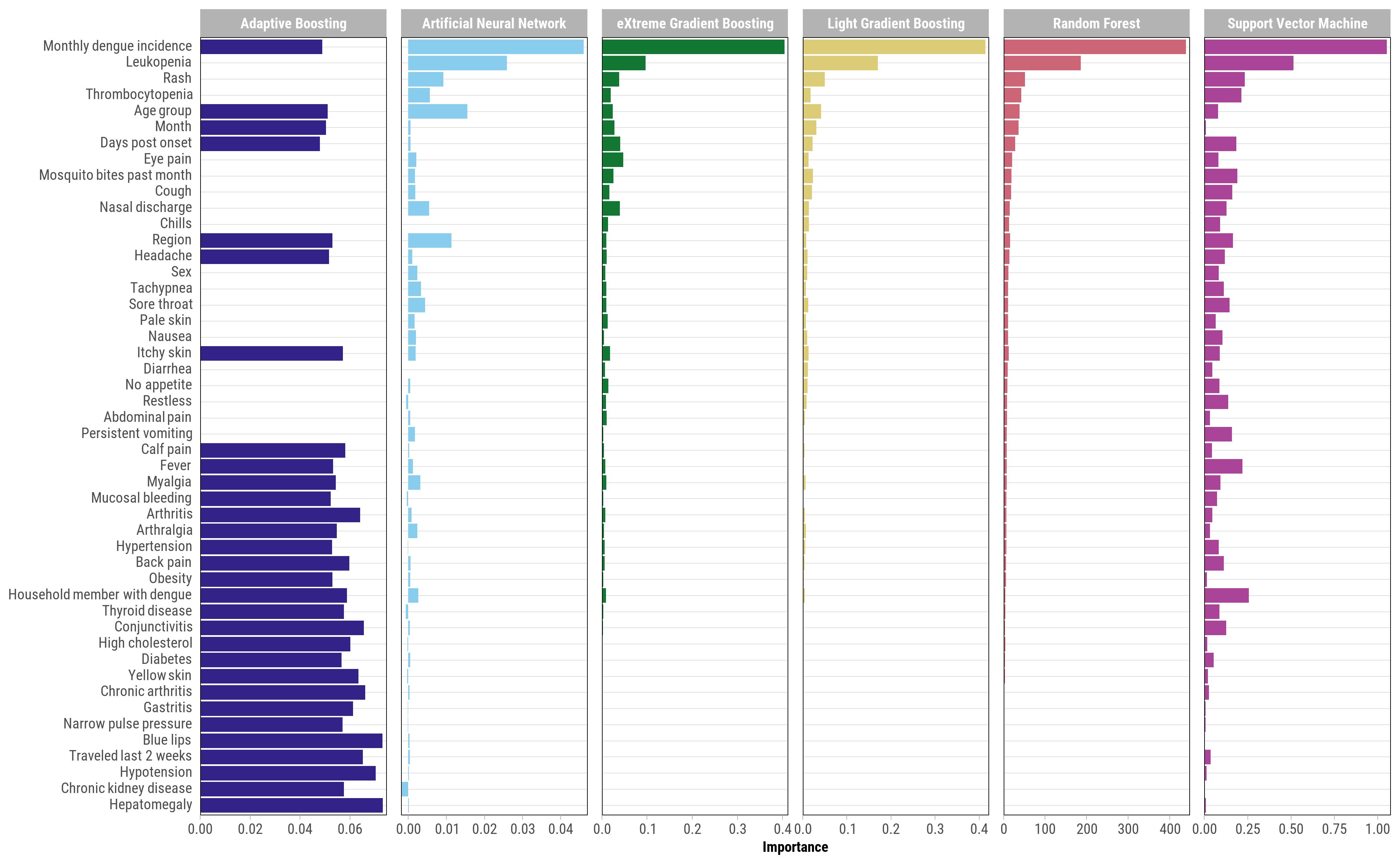

**S5 Fig**. Incremental improvement in AUC by sequentially adding features identified from XGBoost models, SEDSS, May 2012–June 2024.

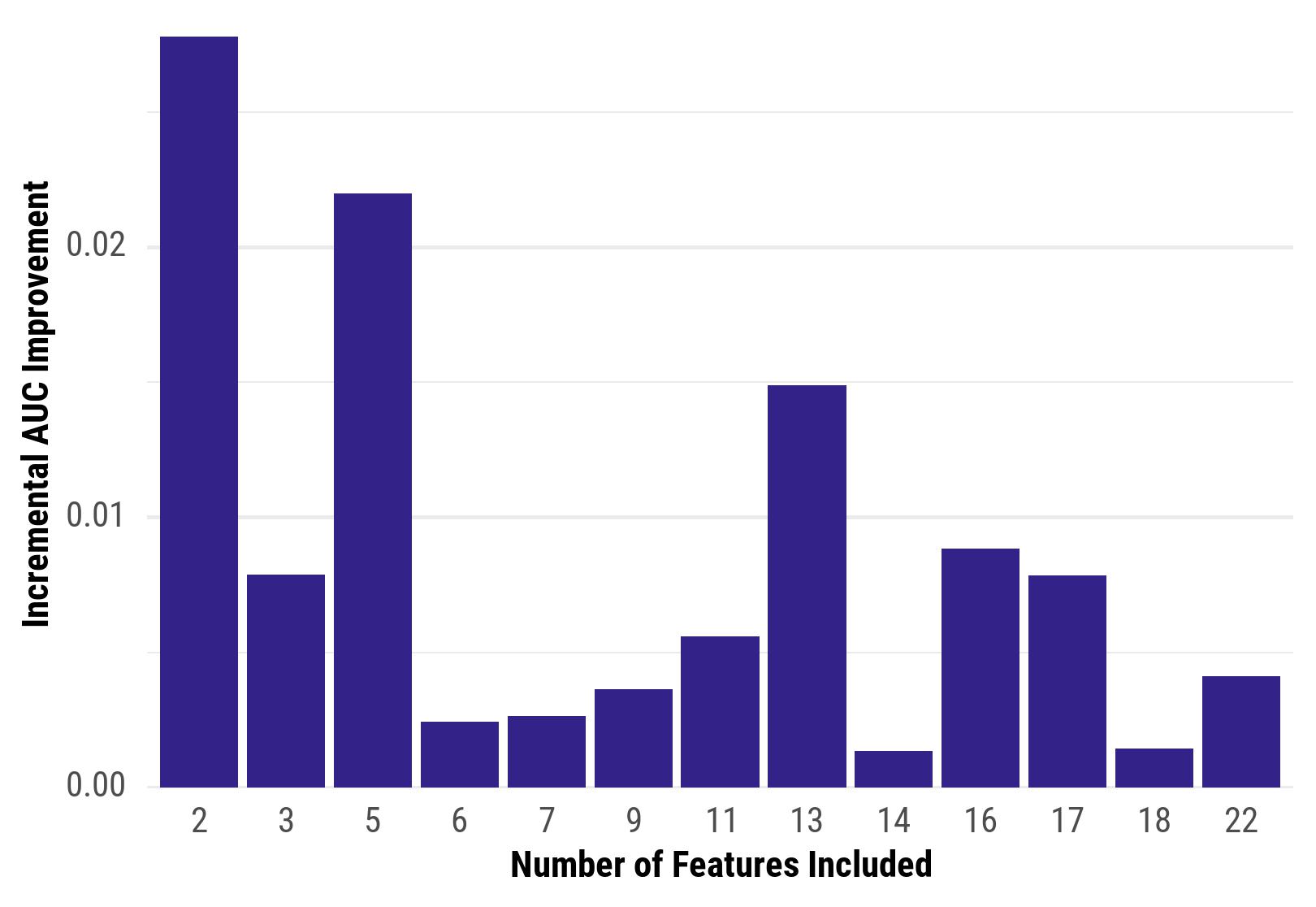
